# SARS-CoV-2 responsive T cell numbers and anti-Spike IgG levels are both associated with protection from COVID-19: A prospective cohort study in keyworkers

**DOI:** 10.1101/2020.11.02.20222778

**Authors:** David Wyllie, Hayley E Jones, Ranya Mulchandani, Adam Trickey, Sian Taylor-Phillips, Tim Brooks, Andre Charlett, AE Ades, EDSAB-HOME investigators, Philippa Moore, John Boyes, Anil Hormis, Neil Todd, Ian Reckless, Andrew Makin, Isabel Oliver

**Author notes:** correspondence:David Wyllie, Public Health England, Forvie Site, Addenbrookes’ Campus, Robinson Way, Cambridge, CB2 0SR.

## Abstract

Immune correlates of protection from COVID-19 are incompletely understood. 2,826 keyworkers had T-SPOT^®^ *Discovery* SARS-CoV-2 tests (measuring interferon-γ secreting, SARS-CoV-2 responsive T cells, Oxford Immunotec Ltd), and anti-Spike S1 domain IgG antibody levels (EuroImmun AG) performed on recruitment into a cohort study. 285/2,826 (10.1%) of participants had positive SARS-CoV-2 RT-PCR tests, predominantly associated with symptomatic illness, during 200 days followup. T cell responses to Spike, Nucleoprotein and Matrix proteins (SNM responses) were detected in some participants at recruitment, as were anti-Spike S1 IgG antibodies; higher levels of both were associated with protection from subsequent SARS-CoV-2 test positivity. In volunteers with moderate antibody responses, who represented 39% (252/654) of those with detectable anti-Spike IgG, protection was partial, and higher with higher circulating T cell SNM responses. SARS-CoV-2 responsive T cell numbers predict protection in individuals with low anti-Spike IgG responses; serology alone underestimates the proportion of the population protected after infection.

## Introduction

The SARS-CoV-2 virus has caused a global pandemic that has killed over a million individuals, disrupted economies, and continues to spread widely; as of March 25, 2021, 4.3 million people have tested positive in the UK alone (1). Disease manifestations vary from asymptomatic or minimally symptomatic infection through to fatal pneumonia (2-5).

Animal data and human epidemiological indicate that immune protection to SARS-CoV-2 diseases can be elicited after natural infection (6-14). However, the relative importance of immune responses elicited following natural infection are not yet clear. Antibodies to SARS-CoV-2 nucleoprotein and spike protein are generated in >90% of cases of symptomatic infection (Table S1) and are associated with clinical protection epidemiologically (6, 7). Potent neutralisation of viral infectivity, predominantly by antibodies directed at the receptor binding domain moiety of the Coronavirus Spike S1 domain, occurs post infection (15), and represent a potential causative explanation for observed associations between serological responses and protection (6, 7). Unfortunately, antibody resistant escape variants can be selected *in vitro* (16), and the recently emergent and highly successful viral variants B.1.1.7 and 501Y.V2 (17) may result from similar *in vivo* selection. In support of this idea, these viruses are less effectively neutralised by antisera from natural infection, or vaccination with Spike protein from, ancestor viruses (18-20). Therefore, an understanding of antibody independent defence mechanisms which might reduce the impact of viral evasion of pre-existing humoral protection is an important topic for investigation.

In asymptomatically infected individuals, generation of SARS-CoV-2 responsive T cells without persistent antibodies has been reported (21-23). Additionally, SARS-CoV-2 responsive T cells have been described in a proportion of the SARS-CoV-2 naive population, likely primed by infection with the endemic common cold *Coronaviridae* (CCCs) (24-27). It has been proposed that such T cells may provide some protection from SARS-CoV-2 infection (27-29).

Reproducible, standardised high throughput serological assays which have minimal cross reactivity with CCCs have been developed (30, 31) and deployed on a large scale, even before a complete picture of their clinical utility was available (32), allowing an understanding of the association between anti-SARS-CoV-2 antibody responses and protection from subsequent COVID-19 (6, 7). By contrast, T cell immune responses to SARS-CoV-2 have been studied in smaller populations using a combination of ELISPOT, flow cytometric analysis of cell surface activation induced markers, as well as intracellular cytokine staining (21-27).

The objectives of this study were i) to describe T cell and antibody responses to the SARS-COV-2 virus in UK keyworkers recruited to the EDSAB-HOME (Evaluating Detection of SARS-CoV-2 AntiBodies at HOME) cohort study (33), and ii) to measure the association between these responses at recruitment and subsequent COVID-19 development.

## Methods

### Study participants

We studied participants in the EDSAB-HOME study (ISRCTN56609224) (33), which recruited and characterised three keyworker “streams” in England. Recruitment occurred from 1 to 26 June 2020 at the end of the UK’s first wave of COVID-19 (Supplementary figure 1). Two streams (Streams A, B) recruited Fire & Rescue or Police service keyworkers, or Health care keyworkers (n=1,139, n=1,533, respectively), independent of any history of COVID-19 disease, asymptomatic SARS-CoV-2 infection, or RT-PCR test results; in Stream C, healthcare workers were purposefully recruited on the basis of a history of prior RT-PCR positive testing (n=154) (Table S2). All participants were administered an identical questionnaire, which asked about demographic details, personal and household illness since 1 Jan 2020, and whether the individual had had RT-PCR tests for SARS-CoV-2. Self-reported SARS-CoV-2 test results were cross-checked against national laboratory records. The cohorts did not include acutely infected individuals; among the 268 (9.4%) cases who had had a positive RT-PCR result prior to recruitment, the test had occurred a median of 63 days prior to recruitment. The cohort would be expected to include both symptomatic and asymptomatically infected individuals, some of whom would not have been identified by the testing then available. On attending a study clinic, participants provided 6ml blood anticoagulated using EDTA used for immunoassays, and 6ml or 10ml lithium heparin anticoagulated blood used for T-SPOT tests.

### Study endpoints

In the period after recruitment, large scale SARS-CoV-2 testing capability was in place in England. For participants, symptom driven nasal/throat RT-PCR testing was available through state and employer routes for those with cough, fever, or disordered taste/smell; all such tests, irrespective of result, are recorded in a national database. Asymptomatic testing was not routinely available, except as part of national surveillance schemes, and in workplace-based settings from 6 November 2020 onward. We followed up participants, defining an endpoint as the first positive SARS-CoV-2 positive RT-PCR test during follow-up, for 200 days to 24 Jan 2021. We obtained details about symptoms and circumstances associated with the positive test from (i) an optional weekly symptom questionnaire sent to volunteers and (ii) a symptom questionnaire sent retrospectively to all individuals with positive tests. We excluded one result obtained from an individual tested as part of national surveillance study of randomly selected asymptomatic individuals, since the positive predictive value of results is much lower in the absence of symptoms(35).

### Disease rates

The daily counts and rates of individuals testing positive for SARS-CoV-2 was obtained from https://coronavirus.data.gov.uk on 26 Jan 2021.

### T-SPOT^®^ *Discovery* SARS-CoV-2 kits

To quantify T cell responses, we used T-SPOT^®^ *Discovery* SARS-CoV-2 kits (T-SPOT hereafter), which use ELISpot technology to detect IFN-γ release from immune cells after exposure to SARS-CoV-2 peptides. This test is similar to the T-SPOT^®^.*TB* test which identifies patients infected with *M. tuberculosis* (34), and has been widely used clinically. Peripheral blood mononuclear cells (PBMCs) were isolated from a whole blood sample using the T-Cell *Select ™* reagent (Oxford Immunotec). After quantification and dilution of recovered cells, 250,000 PBMCs were plated into each well of a T-SPOT^®^ *Discovery* SARS-CoV-2 (Oxford Immunotec) kit. The kit is designed to measure responses to six different but overlapping peptides pools to cover protein sequences of six different SARS-CoV-2 antigens and includes negative and positive controls (Table S3). Peptide sequences that showed high homology to endemic coronaviruses were removed from the sequences, but sequences that may have homology to SARS-CoV-1 were retained. Cells were incubated overnight and interferon-γ secreting T cells detected. To analyse the T-SPOT result of individuals, we computed the sum of T-SPOT responses to groups of panels, subtracting the background (peptides absent) counts (Table S3).

### Laboratory Immunoassays

Our primary serological assay was a commercial immunoassay (EUROIMMUN Ag) measuring IgG against the SARS-CoV-2 Spike protein S1 domain (33). We report results in four categories, referred to as seronegative, weak reactivity, low level, and high level seropositives, depending on the immunoassay signal obtained (see Table S4). We also measured total anti-Nucleoprotein antibody (Roche Elecsys), considering positive assays with index >1.0, which is the manufacturer’s recommended cut-off, as a secondary assay. To assess background levels in these assays, we analysed pre-pandemic plasma from the COMPARE study (36) using the same assays.

### Masking

None of the individuals who ran the laboratory immunoassays had access to any information about the samples. Participants were notified of their EUROIMMUN serological results approximately one month after their clinic visit, with a warning this was not indicative of protection from disease. Participants were not informed of their T-SPOT results.

### Ethics

EDSAB-HOME study was approved by NHS Research Ethics Committee (Health Research Authority, IRAS 284980) on 02-Jun-2020 and PHE Research Ethics and Governance Group (REGG, NR0198) on 21-May-2020. All participants gave written informed consent.

### Statistics

#### Objective 1: Describe baseline immunological responses to the SARS-CoV-2 virus

We displayed T-SPOT counts to six pools of peptides and immunoassay results) as heatmaps generated by hierarchical clustering. We used also depicted correlations between immunological metrics and clinical meta data using heatmaps. We compared T-SPOT responses in individuals at low versus high risk of past infection (see Figure S6 for definitions) using receiver-operator analysis.

#### Objective 2: Measure associations between baseline immunological responses and subsequent COVID-19

To determine whether immunological predictors were associated with protection from COVID-19, we started follow-up 12 days after the last recruitment (i.e. 8 July 2020), the delay being to avoid inclusion of individuals who may have been incubating infection at recruitment. We explored associations between baseline T-SPOT responses and occupational or other risk factors for COVID-19 exposure, and with self-reported COVID-19 associated symptoms. We also quantified associations between the two exposure variables (antibody index and T-SPOT response) and subsequent COVID-19 using Poisson regression, first with categorised exposure variables and, second, estimating continuous relationships following square root transformation of exposure variables. The process by which the transformation was selected is described in Supplementary materials; in sensitivity analyses, we also used logarithmic transformation. Both univariable and multivariable regressions were performed, where multivariable models were adjusted for age as a continuous variable, gender, high risk occupation (defined as being medical or nursing hospital staff), ethnicity (white vs. non-white) as well as the average weekly incidence (individuals testing positive in the last 7 days) rates in the NHS region of residence of the participant. For continuous exposures, we fitted models in sequence: (i) EuroImmun anti-Spike IgG assay result, (ii) T-SPOT response, (iii) both IgG result and T-SPOT response, (iv) both responses and an interaction between them. More detail is given in the Supplementary Methods.

## Results

### Cohort studied

2,867 key workers participated in the study (Fig. S1). 2,826 (98.5%) participants had T-SPOT (interferon gamma release assay) and serological assays performed successfully (Fig. 1). Causes of failure of these assays were raised T-SPOT background in 14/2,867 (0.5%) of cases, with logistical and sample size issues responsible for the other 27 failures (Fig. 1).

**Figure 1.**
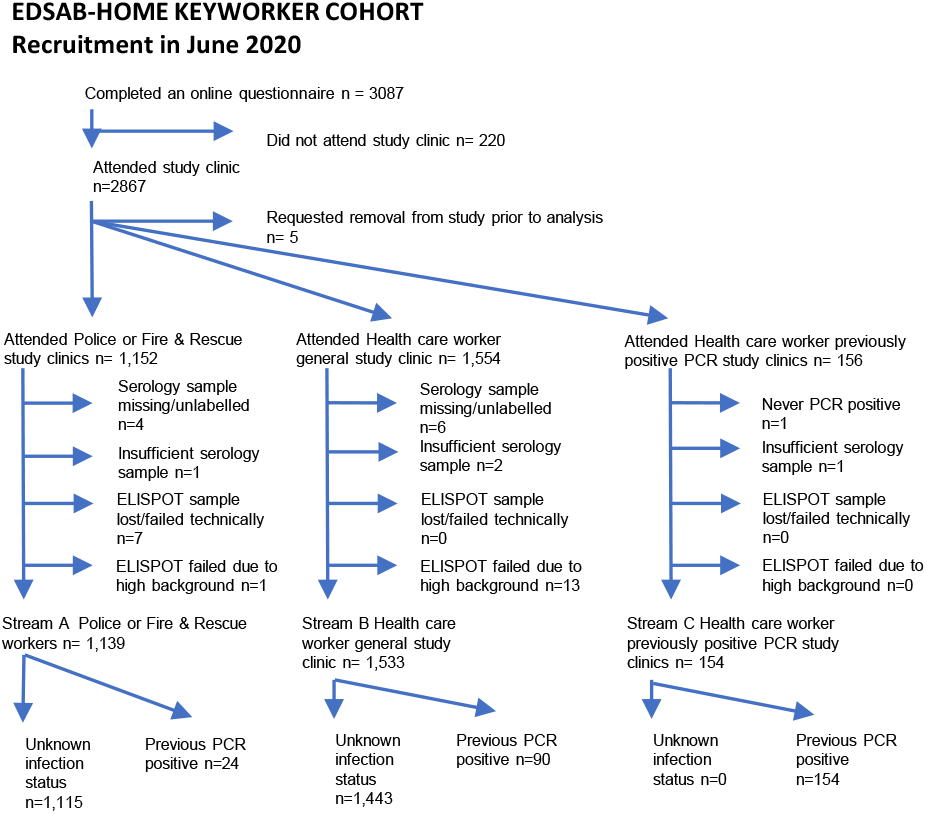
Flow chart illustrating participant flows in the EDSAB-HOME project.

### Evidence of past infection

Characteristics of the cohort, who were working age volunteers from the Fire & Police services during the first wave of the pandemic in the UK from March to June 2020, are shown in Table S2.

Overall, 562/2,826 (15.6%) of participants were seropositive for anti-Spike S1 domain IgG assay using the manufacturer’s cut-off of 1.1 (Fig. S2). An additional 92/2826 (3.3%) individuals had weakly reactive sera (index between 0.35 and 1.1, Fig. S2), of whom 44/92 (48%) also had elevated anti-Nucleoprotein total antibody reactivity. In contrast, among 229 prepandemic samples tested with the same assay, 4/229 (1.7%) had weakly reactive sera none had detectable anti-Nucleoprotein responses (Fig. S2B). This suggested that some study participants with weak reactivity may have been previously infected with SARS-CoV-2. In support of this, we observed a higher proportion of individuals with previous SARS-CoV-2 RT-PCR test positivity among study participants with weak reactivity (17/92, 18.5%) vs. seronegative (6/2172, 0.3%) participants.

### Concordant T cell responses against multiple SARS-CoV-2 peptide pools

A wide range of T cell responses to peptide pools from SARS-CoV-2 proteins (Table S3) were detected using the T-SPOT test (Figure 2). We noted strong positive correlations between T-SPOT responses to Spike S1, S2 domains, Membrane, Nucleoprotein (r ∼= 0.6) (Figure S3) and, therefore, we analysed the background-subtracted sum of T-SPOT responses to Spike, Membrane and Nucleoprotein (T-SPOT SNM results, see Table S3 for details). We also separately analysed the background subtracted sum of T-SPOT Envelope and Structural protein responses (T-SPOT ES results, Figure 2). We observed that T-SPOT SNM responses were markedly stronger than ES responses (Figure 3; for a comparison with anti-Nucleoprotein total antibody see Figure S4).

**Figure 2.**
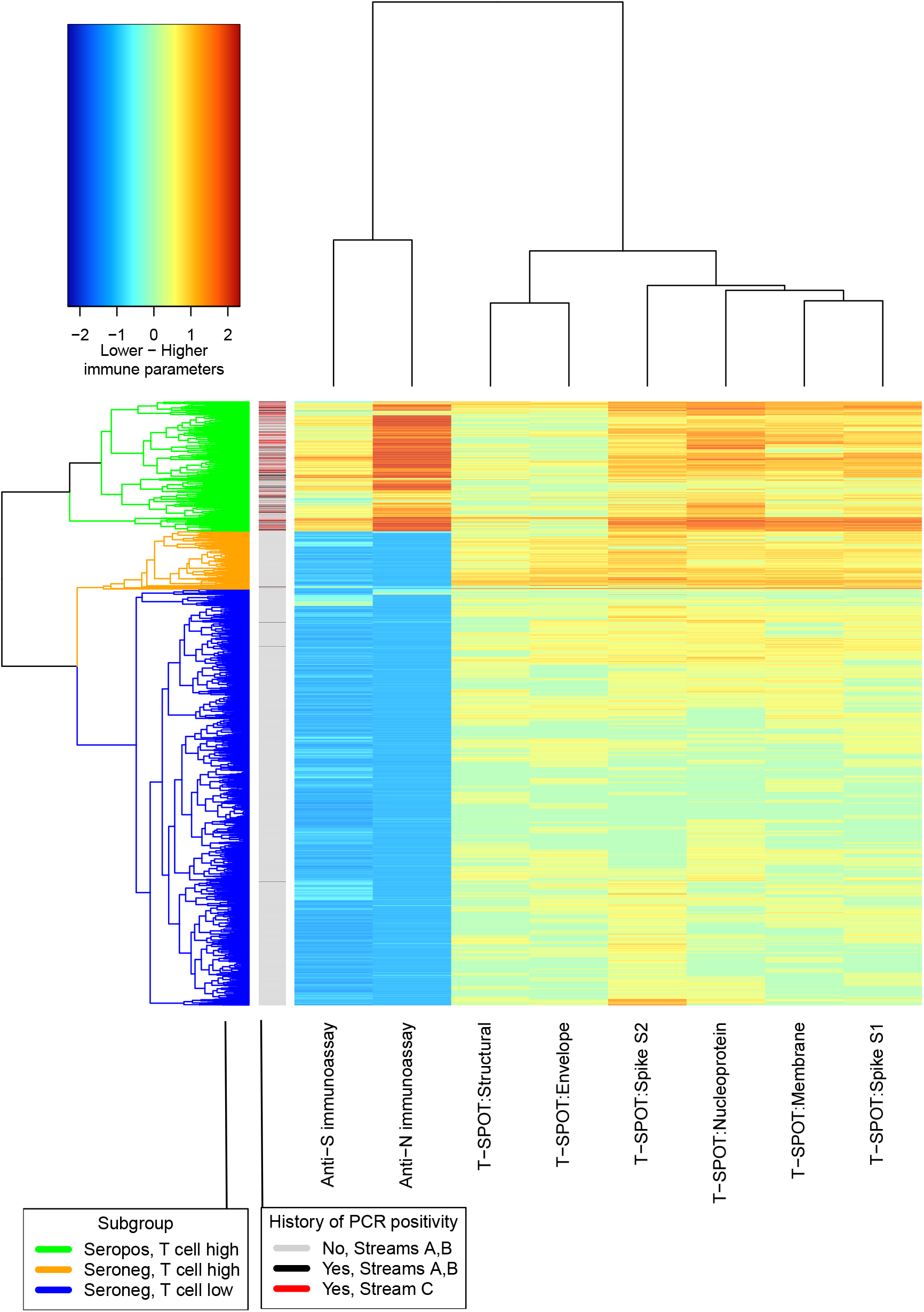
Hierarchical clustering of responses to Spike S1 domain, Spike S2 domain, Nucleoprotein, Membrane protein, structural proteins and Envelope proteins, as well as anti-Nucleoprotein and S1 serological responses in 2,826 unselected keyworkers from streams A,B and C. Footnote: Data were log-transformed prior to clustering; units are arbitrary. The previous PCR positivity status of the participants is shown in a guide bar on the left of the main heatmap.

**Figure 3.**
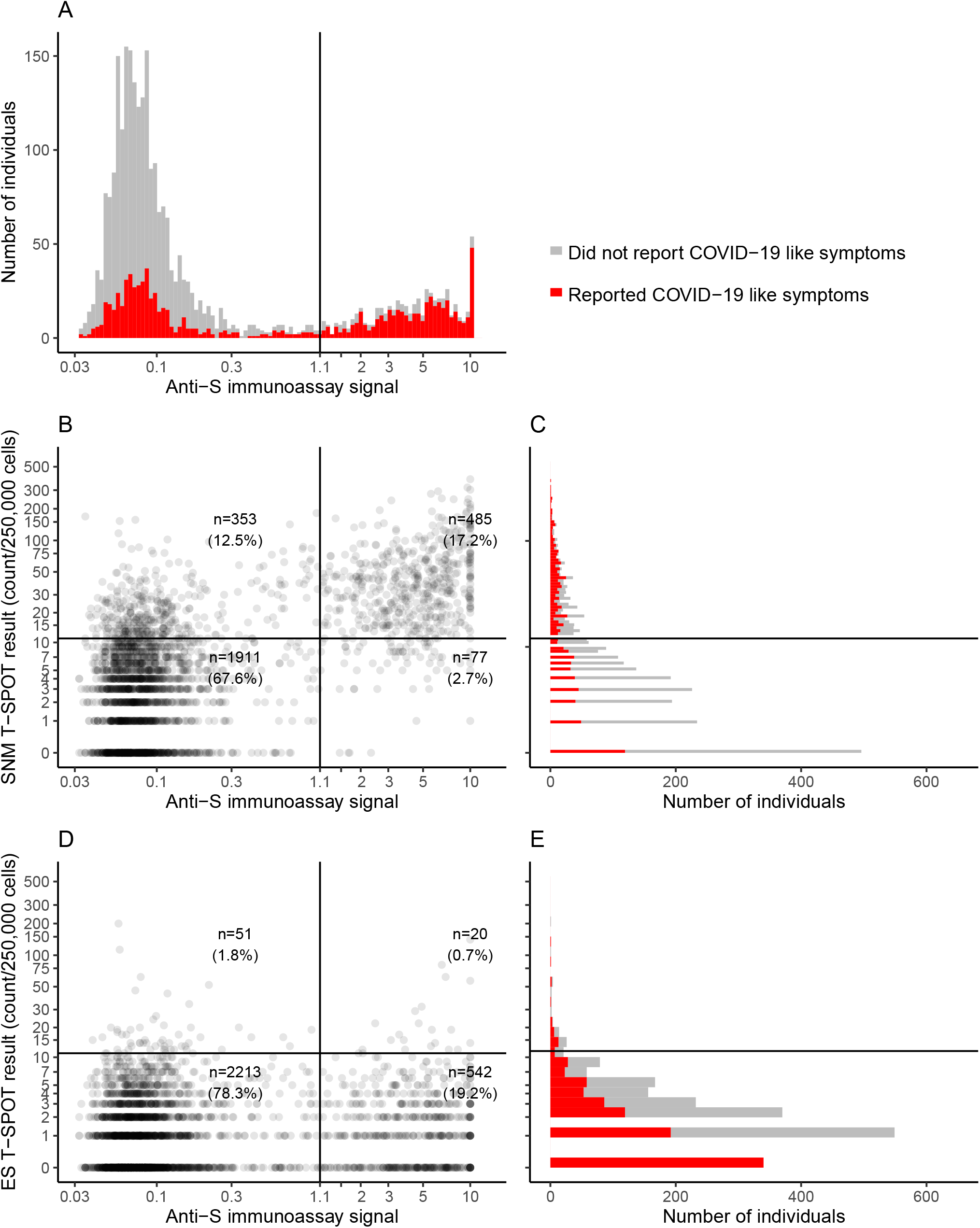
SNM responsive T cells responses and their relationship to anti-S1 IgG serological responses in 2,826 individuals (A) distribution of anti-Spike S1 IgG antibody responses (EUROIMMUN), and its relationship to symptoms. The vertical line is at 0.8, a manufacturer specified cutoff. (B) bivariate plots of anti-S1 IgG responses (EUROIMMUN) and the sum of Spike, Nucleoprotein and Membrane protein responsive T cell numbers. The horizontal line corresponds to 12 spots / 250,000 cells. (C) distribution of the sum of Spike, Nucleoprotein and Membrane protein responsive T cell numbers. (D) As in B, but for Envelope and structural protein responsive T cell numbers; (E) Distribution of Envelope and structural protein responsive T cell numbers. In marginal histograms (A,C,E), the number of individuals reporting symptoms is depicted in red.

### T-SPOT SNM responses and anti-Spike IgG following SARS-CoV-2 infection

Among the 268/2826 individuals with confirmed previous SARS-CoV-2 infection, evidenced by a historical positive RT-PCR test, median T-SPOT SNM responses were 40 (interquartile range [IQR]: 22-67; 5^th^, 95^th^ centiles 8-167 cells per 250,000 PBMC) (Table S5). Comparing individuals with previous SARS-CoV-2 RT-PCR positivity with a subset of the cohort stringently selected to have a low risk of past SARS-CoV-2 infection (Figure S6), T-SPOT SNM results differentiate these two populations with area under curves of 0.96 and an optimal differentiation point of 12 cells / 250,000 PBMC (Figure S7A; T-SPOT ES results do not have this property, Fig S7B). However, there is overlap in T-SPOT SNM responses in individuals with confirmed disease and individuals we selected to be at low risk of past infection (Fig. S6).

### Associations between serostatus, T-SPOT SNM responses, and COVID-19 risk factors and symptoms

While T-SPOT SNM results were associated with multiple well-characterised COVID-19 features, including fever, muscle aching, fatigue, headache, with abnormal sense of taste and smell in both the individual and in their household, and with occupation in the cohort overall (Figure 4A, Table S6), none of these associations were observed among the subset of seronegative individuals (Table S6).

**Figure 4.**
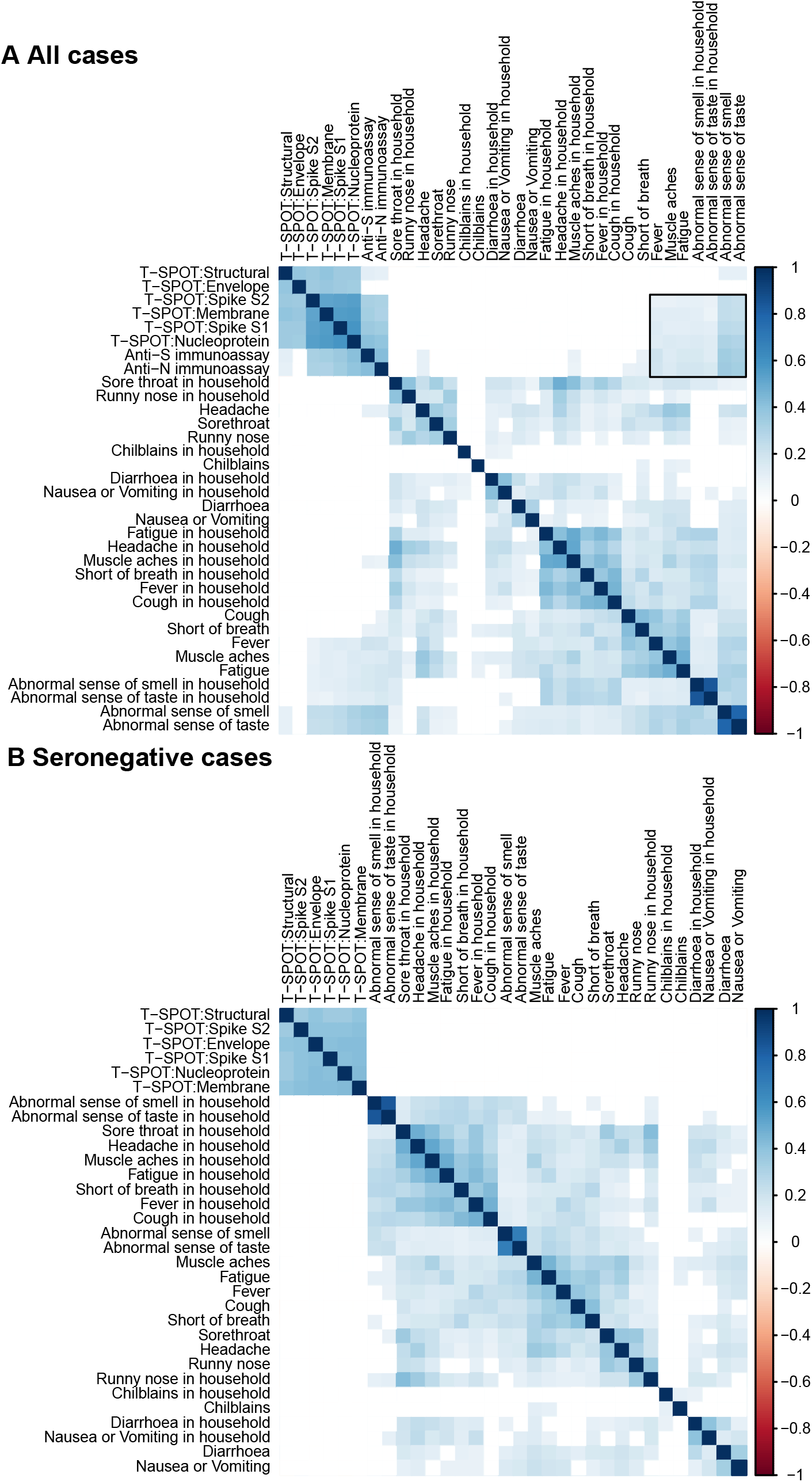
Correlation matrices showing the relationship between individual or household self-reported symptoms, T cell numbers and immunoassay results in (A) 2,672 individuals from Streams A,B (B) 2,197 seronegative individuals. Footnote: Clustering was performed independently in the two datasets. The order of the columns minimises differences between individuals. Only strong correlations are shown. Colour scale reflects Spearman’s correlation coefficient, ρ. In (A), the box denotes a correlation between various immune parameters and symptoms. Comparable correlations are not seen if symptomatic individuals are excluded (B). Serological data is not included in (B) as in (B) all individuals are seronegative.

The only exception was age. In seronegative individuals, T-SPOT counts fell with age (e.g. median: 5; 5^th^, 95^th^ centiles 0-28.5 cells/250,000 in 18-25 year olds vs. 3; 0-15 cells /250,000 in >60 year olds. By contrast, in seropositive individuals, counts rose with increasing age (median, 19.5; 95^th^ centile 4.95, 100.5 in 18-25 year-olds vs. 45; 8.6, 141.6 in over 60s) (see also Table S6).

Thus, in seronegative individuals, T-SPOT counts decline with age and are not associated with past infection, with the converse true in seropositive individuals.

### Positive SARS-CoV-2 tests and symptoms

Overall, 285/2826 (10.1%) of participants had positive SARS-CoV-2 tests during a 200 day follow up (Table 1). At least 75% (216/285) were symptomatic: in 191 cases, questionnaires reporting symptoms were returned, while 25 positive results occurred before any asymptomatic testing was available, although questionnaires were not returned. Only 15/216 (7%) of respondents reported having no symptoms (as listed in Table S6) when they were tested. In 54 cases (18%), questionnaires were not returned, and so symptom status is unknown. Of 191 symptomatic respondents, symptoms reported included shortness of breath (23%), cough (33%), fever (33%), sore throat (33%), runny nose (31%), headache (53%), muscle aching (53%), abnormal taste / smell (36%), fatigue (77%), and diarrhoea (17%).

**Table 1.**
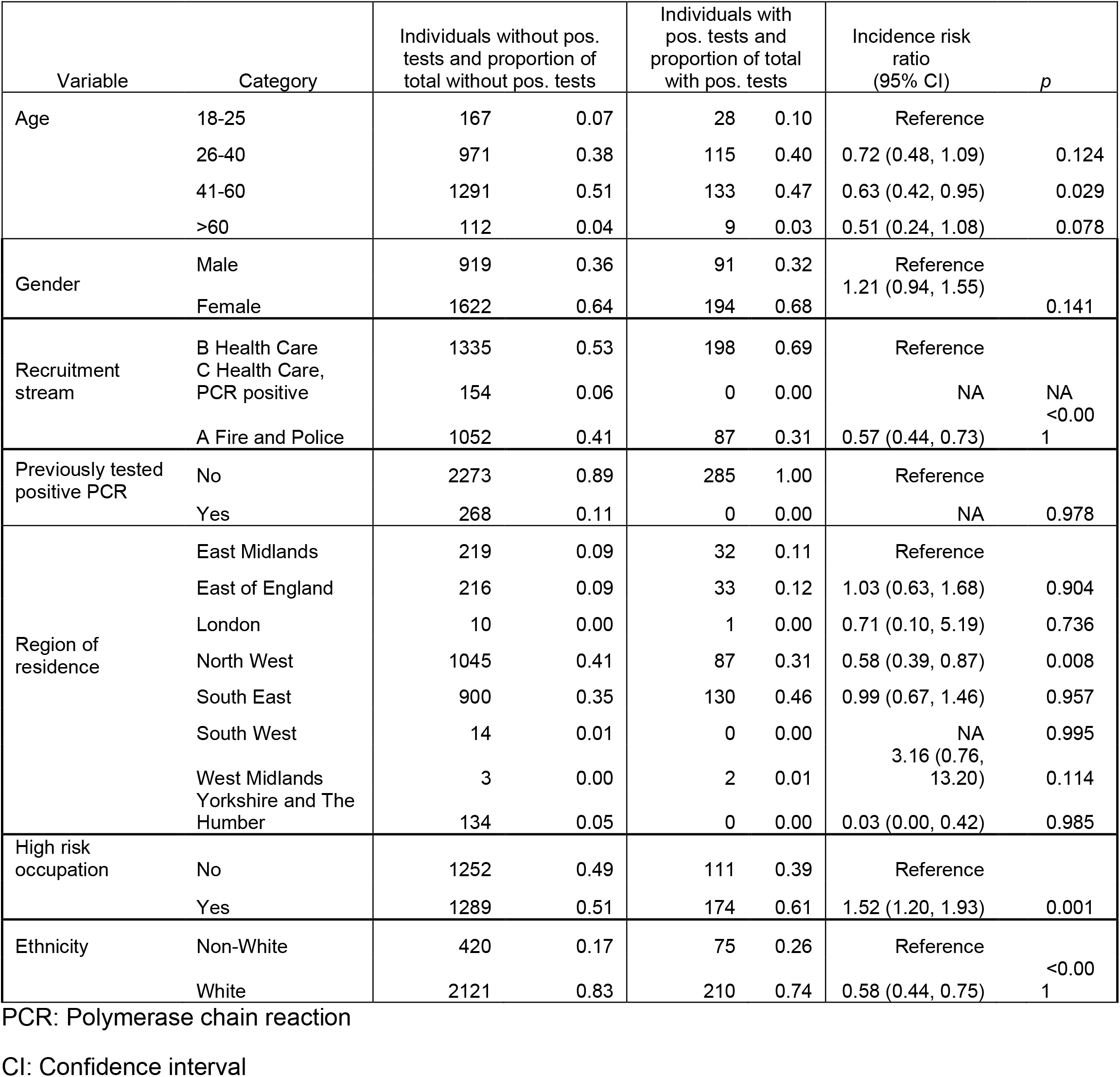
Risk factors for testing PCR Positive on follow-up

### Associations between anti-Spike IgG index and T-SPOT SNM responses and subsequent disease

The risk of testing positive for SARS-CoV-2 fell as both anti-Spike IgG index and T-SPOT SNM responses increased (Figures 5, 6, Tables 2-5). These associations were robust to adjustment for age, gender, region of residence, high risk occupation, ethnicity, and background infection rates (Tables 3-5).

**Figure 5.**
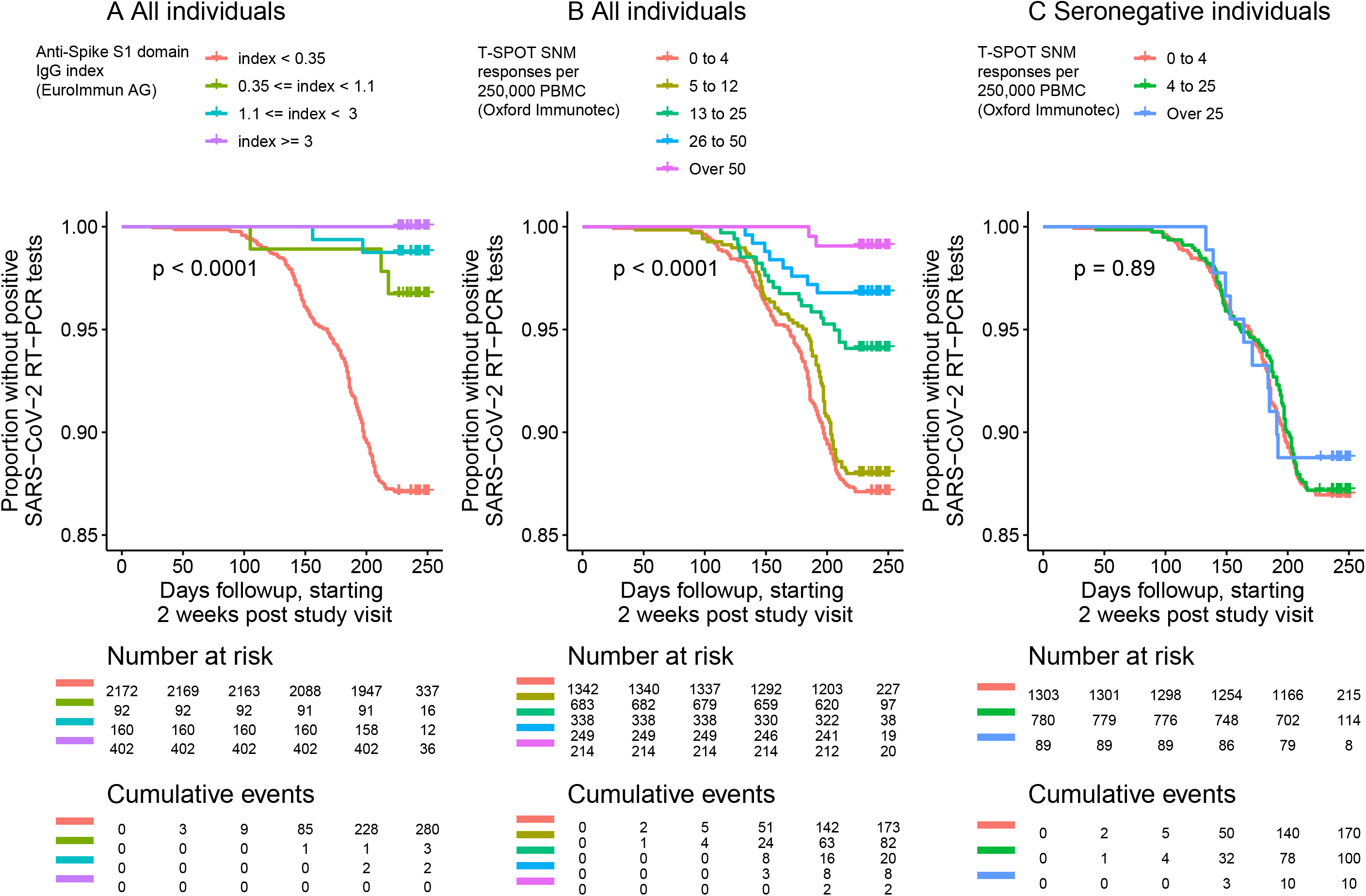
Kaplan-Meier survival curves in showing time to testing positive for SARS-CoV-2, A) stratified by IgG responses against Spike protein for all individuals, B) stratified by T-SPOT SNM responses for all individuals, and C) stratified by T-SPOT SNM responses for seronegative individuals*.. *Seronegative: EuroImmun anti-S IgG index < 0.35.

**Figure 6.**
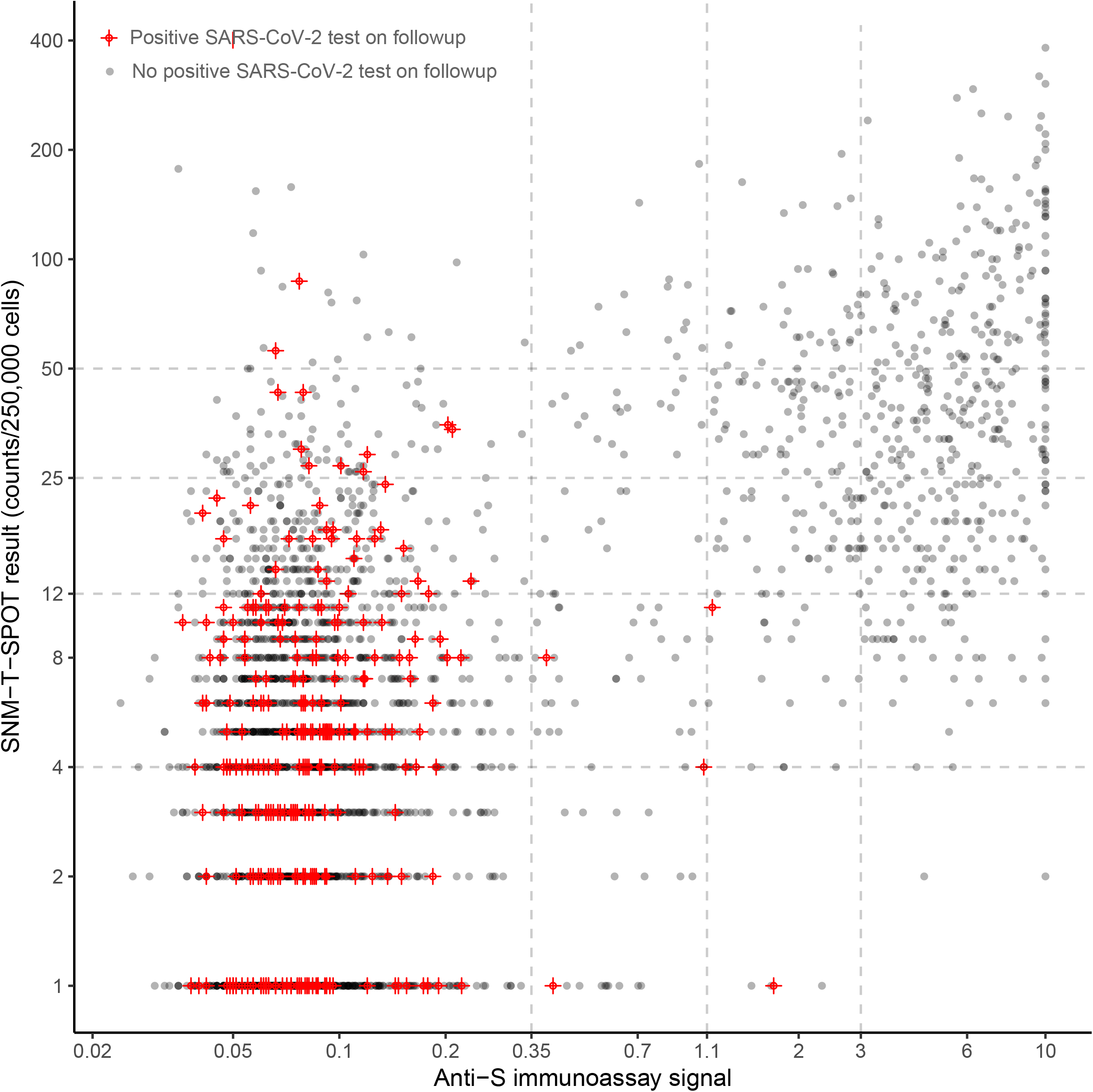
Anti-Spike immunoassay results and T-SPOT SNM responses, illustrating the immunological metrics of those with positive SARS-CoV-2 tests, and those without. Footnote: Dotted vertical lines reflect the margins of the categorisation presented in Table 2.

**Table 2.** Percentage of individuals testing positive for SARS-CoV-2, stratified by immunological markers

| T-SPOT response to SNM pools (per 250,000 cells) | Anti-Spike immunoassay signal (EuroImmun index) * |  |  |  | Total |
| --- | --- | --- | --- | --- | --- |
|  | Seronegative<br>0 to 0.35 | Weakly<br>reactive<br>0.35 to 1.1** | Low level<br>seropositive<br>1.1 to 3 | High level<br>seropositive Index<br>>3 |  |
| **0-4** | 13% (170/1303) | 7% (2/26) | 12% (1/8) | 0% (0/5) | 13% (173/1342) |
| **5-12** | 13% (80/587) | 3% (1/26) | 5% (1/19) | 0% (0/51) | 11% (81/683) |
| **13-25** | 11% (20/193) | 0% (0/8) | 0% (0/47) | 0% (0/90) | 6% (20/338) |
| **26-50** | 11% (8/69) | 0% (0/17) | 0% (0/53) | 0% (0/110) | 3% (8/249) |
| **>50** | 10% (2/20) | 0% (0/15) | 0% (0/33) | 0% (0/146) | 1% (2/214) |
| **Total** | 12% (280/2172) | 3% (3/92) | 1% (2/160) | 0% (0/402) | 10% (285/2826) |

\* The manufacturer's cutoff is 1.1 for 'confirmed positive' cases. \*\* Antibody ranges are closed on the left, so an interval of 0 to 0.35 denotes [0,0.35) and would include 0 but not 0.35. See Table S4 for definitions.

**Table 3.** Rates and rate ratios (RR) of testing positive for SARS-CoV-2, stratified by T-SPOT SNM response

|  | **T-SPOT SNM response categories** |  |  |  |  |
| --- | --- | --- | --- | --- | --- |
|  | **0-4** | **5-12** | **13-25** | **26-50** | **>50** |
| Percentage of individuals testing positive for SARS-CoV-2 | 13% (173/1342) | 11% (81/683) | 6% (20/338) | 3% (8/249) | 1% (2/214) |
| Rate per 100,000 person days | 60.8 (52.4, 70.5) | 56.4 (45.3, 70.0) | 27.7 (17.9, 42.9) | 15.1 (7.5, 30.2) | 4.4 (1.1, 17.6) |
|  | **Relative risk of testing positive (rate ratio with 95% confidence intervals)** |  |  |  |  |
| Unadjusted RR | Reference | 0.93 (0.71-1.21) | 0.46 (0.29-0.72) | 0.25 (0.12-0.50) | 0.07 (0.02-0.29) |
| Adjusted RR* | Reference | 0.91 (0.70-1.18) | 0.43 (0.27-0.68) | 0.24 (0.12-0.50) | 0.08 (0.02-0.32) |
| Additionally adjusted for background rates RR** | Reference | 0.90 (0.69-1.18) | 0.43 (0.27-0.68) | 0.25 (0.13-0.52) | 0.09 (0.02-0.36) |

\* Adjusted for sex, ethnicity, age (continuous), high-risk occupation, region.
\*\* As above but additionally adjusted for time-updated regional SARS-CoV-2 infection rates

In strongly seropositive individuals (n=402), no events were observed at any level of SNM T cell responses (Table 2). By contrast, among 2,172 individuals who were seronegative, 12.9% (285) tested positive over the follow-up period. Among the 252 individuals with weak reactivity or low level seropositivity (antibody index 0.35 to 3), 5 individuals developed events (2.0%) (Table 4). In this group, protection was associated with T-SPOT SNM results: 5/79 of those with 0-12 SNM responsive cells had positive PCR tests on follow-up, vs. 0/173 with more than 12 SNM responsive cells (Table 2). A post hoc Wilcoxon test provided evidence that higher SNM T cell responses were associated with enhanced protection within the subgroup with antibody index between 0.35 and 3 (*p*=3×10^−3^). This association with SNM T cell responses was not observed in the seronegative subjects (p = 0.47, Wilcoxon test).

**Table 4.** Rates and rate ratios (RR) of testing positive for SARS-CoV-2, stratified by anti-Spike antibodies

|  | Anti-spike antibody categories |  |  |  |
| --- | --- | --- | --- | --- |
| | Seronegative<br>0 to 0.35 | Weakly reactive<br>0.35 to 1.1<sup>\$</sup> | Low level seropositive<br>1.1 to 3 | High level<br>seropositive Index<br>>3 |
| Percentage of individuals testing positive for SARS-CoV-2 | 13% (173/1342) | 11% (81/683) | 6% (20/338) | 3% (8/249) |
| Rate per 100,000 person days (95% confidence intervals) | 60.7 (54.0, 68.3) | 15.1 (4.9, 46.9) | 5.9 (1.5, 23.6) | 0 |
|  | **Relative risk of testing positive (rate ratio with 95% confidence intervals)** |  |  |  |
| Unadjusted RR | Reference | 0.25 (0.08-0.78) | 0.10 (0.02-0.39) | 0 |
| Adjusted RR* | Reference | 0.24 (0.08-0.75) | 0.08 (0.02-0.34) | 0 |
| Additionally adjusted for background rates RR*** | Reference | 0.24 (0.08-0.75) | 0.09 (0.02-0.38) | 0 |
| Additionally adjusted for background rates and T-cell categories RR**** | Reference | 0.27 (0.08-0.85) | 0.12 (0.03-0.49) | 0 |

\* The manufacturer's cut-off is 1.1 for 'confirmed positive' cases. See Table S4 for definitions.
\$ Antibody ranges are closed on the left, so 0 to 0.35 denotes [0,0.35] i.e. would include 0 but not 0.35. See Table S4 for definitions
\*\* Adjusted for sex, ethnicity, age (continuous), high-risk occupation, region.
\*\*\* As above but additionally adjusted for time-updated regional SARS-CoV-2 infection rates
\*\*\*\* As above but also adjusted for T-cell count categories

When modelling continuous relationships, we found support for an interaction between SNM T-SPOT responses and anti-Spike antibody levels in protection from SARS-CoV-2 infection in the cohort studied (Table 5). Sensitivity analyses modelling log, rather than square root, transformed exposure variables similarly found evidence of interaction (Table S7). Put alternatively, modelling approaches indicated that (as is suggested by inspection of the raw data, Figure 6, and stratified analyses, Tables 2-4), disease protection increases as both SNM T cell responses and antibody levels increase.

**Table 5:** Incidence rate ratios (95% confidence intervals) of testing positive for SARS-CoV-2, modelling immunological metrics as transformed continuous variables

|  | **Model 1:**<br>**antibodies only** |  | **Model 2:**<br>**T cells only** |  | **Model 3:**<br>**antibodies and T cells** |  | **Model 4:**<br>**antibodies, T cells, and**<br>**interaction term** |  |
| --- | --- | --- | --- | --- | --- | --- | --- | --- |
|  | **Unadjusted** | **Adjusted\*** | **Unadjusted** | **Adjusted\*** | **Unadjusted** | **Adjusted\*** | **Unadjusted** | **Adjusted\*** |
|  | **Incidence rate ratios (95% confidence intervals)** |  |  |  |  |  |  |  |
| Square-root transformation of Anti-Spike IgG antibody index | 0.09<br>(0.03, 0.26) | 0.08<br>(0.03, 0.23) | N/A | N/A | 0.09<br>(0.03, 0.28) | 0.08<br>(0.03, 0.25) | 1.80<br>(0.28, 11.50) | 1.94<br>(0.29, 12.74) |
| Square-root transformation of T-SPOT response to SNM pools | N/A | N/A | 0.90<br>(0.87, 0.93) | 0.89<br>(0.86, 0.92) | 0.99<br>(0.95, 1.03) | 0.99<br>(0.95, 1.03) | 0.99<br>(0.96, 1.04) | 1.00<br>(0.96, 1.04) |
| Interaction term between the two variables above | N/A | N/A | N/A | N/A | N/A | N/A | 0.60<br>(0.38, 0.96) | 0.58<br>(0.36, 0.93) |
| *p*-value of model fit vs Model 1 | N/A | N/A | N/A | N/A | 0.4735 | 0.4995 | 0.0171 | 0.0127 |
| *p*-value of model fit vs Model 3 | N/A | N/A | N/A | N/A | N/A | N/A | 0.0058 | 0.0040 |

Adjusted for age, gender, high-risk occupation, and white ethnicity.

## Discussion

In a large, prospective cohort study of keyworkers recruited in June 2020, about 16% of participants had clear serological evidence of past SARS-CoV-2 infection. We measured two immune correlates of protection against COVID-19 using commercial immunoassays: anti-Spike IgG antibodies (measured using kits from EuroImmun AG) and SARS-CoV-2 responsive T cell numbers (T-SPOT Coronavirus Discovery Kits, from Oxford Immunotec Plc). Using the T-SPOT assay, which is designed to minimise cross-reactivity with other similar viruses, T-cells responsive to peptides from several SARS-CoV-2 antigens were detected; responses to Spike, Nucleoprotein and Membrane proteins (SNM responses) were co-correlated, and coexisted with anti-Spike and anti-Nucleoprotein antibodies individuals with a history of microbiologically confirmed infection. This result is compatible with smaller studies in healthcare workers studying T cell assays using intracellular cytokine staining (21-23). By contrast to responses to SNM proteins, T cell responses to envelope and various structural proteins were much weaker and were not increased in those with a history of SARS-CoV-2 infection vs. those who had not.

We observed that higher anti-Spike antibody levels and SARS-CoV-2 responsive T cell numbers are associated with protection from testing positive for SARS-CoV-2; our data suggest an interaction between antibody levels and circulating SARS-CoV-2 responsive T cell numbers, such that, among individuals with modest antibody levels (EuroImmun anti-Spike IgG assay index 0.35 to 3, levels seen in about 40% of individuals with detectable antibody in our study) subsequent COVID-19 rates were lowest in those with higher T cell counts. This suggests a role for T cells in SARS-CoV-2 control, which is biologically plausible via multiple mechanisms, including direct T cell mediated virus control, T cell help for subsequent B cell antibody production, and as a marker for a memory B cell population primed by natural infection (40).

Interestingly, although the highest SNM responses were seen in individuals who also had detectable anti-Spike IgG antibody, in some individuals SNM responses comparable with those observed in SARS-CoV-2 survivors were seen in the absence of anti-Spike IgG. This remained true even if a very stringent cut-off was used for seropositivity, such as EuroImmun assay index > 0.35, which is much lower than the manufacturer’s cut-off of 1.1 for a confirmed positive. It is possible that this is the case because some individuals, after exposure to SARS-CoV-2, retain circulating T cell responses when serological responses decline to undetectable levels, or never make antibody at all(21-23). However, our epidemiological data do not support this explanation: absent detectable anti-Spike IgG, T-SPOT SNM responses were not associated with either occupational risk of COVID-19 (e.g. occupation) nor with a history of canonical COVID-19 symptoms. Although we cannot exclude previous SARS-CoV-2 infection without persistent antibody in a proportion of cases, exposure to other viruses, perhaps Common Cold Coronaviruses (CCCs) (24-27) inducing T cells cross-reactive with SARS-CoV-2 in the absence of persistent serological cross-reactivity(21, 22) is another possibility. Independent of the mechanism involved, it is important to note that, absent antibody against SARS-CoV-2, such cells may offer little SARS-CoV-2 protection since among seronegative volunteers we did not detect significant disease protection associated with T-SPOT SNM responses.

The size of the study, its cohort design, the ascertainment of endpoints using a standardised, national infrastructure are strengths of the study, and the collection of anti-SARS-CoV-2 serological responses and T cell responses at recruitment allow us to go beyond previous cohort studies that have examined associations between antibody levels and subsequent disease only(6, 7). Limitations include our measurement of immune correlates of protection an average of two months after infection (among those with PCR confirmation); we also counted circulating SARS-CoV-2 responsive interferon-γ secreting cells, but did not phenotype them (24), and so may have missed prognostic immunophenotypes in circulating cells, or aspects of their kinetics. In our endpoints, we did not consider recurrences, nor did we measure disease severity: immunological correlates of protection from severe disease, and from recurrence, might differ from those ascertained here.

Given the observational nature of the study, confounders might have affected the associations we observed. Multivariable modelling has been used to address this, but some factors cannot be controlled readily. For example, participants knew their symptom history and antibody status, so seropositive or previously symptomatic participants may have been more likely to allow themselves to be exposed to SARS-CoV-2. Secondly, some individuals may have been infected without being tested (and acquired protection) subsequent to the clinic visits. Thirdly, individuals at the highest risk of occupational exposure on follow-up may have been at highest risk of having been infected prior to recruitment. However, all these potential biases would be expected to dilute any immunology - protection association.

Overall, this study suggests that clinical serodiagnostics with performance similar to the assay used here may underestimate the population with SARS-CoV-2 immunity for two reasons. Firstly, protection is evident at low antibody concentrations below the manufacturer’s assay cut-off – so, if a clinical laboratory was reporting using this cut-off, a proportion of individuals with negative results would have low levels of antibodies and some degree of protection. Secondly, Coronavirus Spike, Nucleoprotein and Membrane response T cell numbers potentiate the protection seen at a given antibody level. This may reflect antibody independent T-cell control of viral replication(40). Such mechanisms, if present, may mitigate the impact of viral mutants less effectively neutralised by antisera induced by vaccination or ancestral strains (17-20). The magnitude of any such effect is of obvious public health importance and deserves additional investigation. The conduct of, and comparisons between, such investigations may be assisted by the availability of standardised T-SPOT kits used in this study, which are of similar format to the widely used T-SPOT®.*TB* tests used for tuberculosis diagnosis(34).

## Data Availability

Data available on reasonable request to corresponding author.

## Data availability

The data analysed is available from the corresponding author on request.

