## Supplementary Material for "SARS-CoV-2 responsive T cell numbers and anti-Spike IgG levels are both associated with protection from COVID-19: A prospective cohort study in keyworkers"

| | |
|---|---|
| Title | SARS-CoV-2-CoV-2 responsive T cell numbers are associated with protection from COVID-19: A prospective cohort study in key workers |
| Authors' names and affiliations | David Wyllie[1+], Ranya Mulchandani[1], Hayley E Jones[2], Sian Taylor-Phillips[3], Tim Brooks[1], Andre Charlett[1], AE Ades[2], EDSAB-HOME investigators, Andrew Makin[4], Isabel Oliver[1]<br>+ correspondence:<br>1 Public Health England<br>2 University of Bristol<br>3 University of Warwick<br>4 Oxford Immunotec Ltd |
| Correspondence | David Wyllie, Public Health England, Forvie Site, Addenbrookes' Campus, Robinson Way, Cambridge, CB2 0SR. |

#### Methods

##### Study participants

We studied participants in the EDSAB-HOME study (1), which recruited three key worker ”streams”. Two streams (Streams A, B) comprised Fire & Rescue or Police service key workers, or Health care key workers (n=1,139, n=1,533 respectively), independent of any history of COVID-19 disease or RT-PCR testing; for Stream C, a history of prior RT-PCR positive testing was required (n=154). The cohorts did not include acutely infected individuals; among the 268 (9.4%) cases who had had a prior positive RT-PCR result, the test had occurred a median of 63 days prior to recruitment. Recruitment occurred in June 2020. All participants provided written informed consent.

Individuals who were currently working at their place of work, or eligible to do so, aged 18 years or over, able to read English and to provide informed consent were eligible to enrol, as described (ISRCTN56609224). Anyone currently experiencing COVID-19 compatible symptoms, who had experienced any in the last 7 days, meeting the UK Government’s criteria for “exceptionally vulnerable”, or taking part in any COVID-19 vaccine trials was ineligible.

Participants completed an online questionnaire prior to clinic attendance. On attending a study clinic, venous blood samples were taken from each participant. Each subject had a 6ml blood sample anticoagulated using EDTA, plasma was separated by centrifugation and stored at -80$^{o}$C until use and 6ml (for the first 500 participants) or 10ml (thereafter) of blood anticoagulated with lithium heparin, stored at room temperature and shipped overnight to Oxford Immunotec (Milton Park, Oxon, UK) for T-SPOT® *Discovery* SARS-CoV-2 tests.

##### Laboratory Immunoassays

We characterised serological responses against SARS-CoV-2 using two commercial immunoassays: Roche Elecsys® anti-SARS-CoV-2 (2), and EUROIMMUN anti-S IgG immunoassays (1). We report the association between protection and the EUROIMMUN assay in this paper. Results failing immunoassays for technical reasons were repeated, as per normal clinical practice, and samples with insufficient volume for testing were excluded from analyses. We considered these assays positive if the Roche Elecsys® immunoassay signal was over 1.0, as recommended by the manufacturer, or the EUROIMMUN immunoassay index was over the ‘confirmed’ cutoff of 1.1 (1).

#### T-SPOT Discovery SARS-CoV-2 Testing

In brief, cells were purified from peripheral blood and SARS-CoV-2 responsive T cell numbers quantitated using a T-SPOT Discovery SARS-CoV-2 tests, which are based on the same proven analytical platform as the T-SPOT®.*TB* (tuberculosis) kit also produced by Oxford Immunotec.

In detail, peripheral blood mononuclear cells (PBMCs) were isolated from a whole blood sample using T-Cell *Select* reagent (Oxford Immunotec). The T-Cell *Select* kit contains a proprietary set of reagents consisting of buffer concentrate, antibodies, and superparamagnetic beads. Diluted T-Cell *Select* buffer is added to the whole blood sample to facilitate cell purification and reduce red blood cell contamination, and then antibodies are added which bind to the requisite immune cells in the sample. Cell separation was done using a validated automated platform, with a KingFisher magnetic separator (ThermoFisher). Cells were resuspended, counted using Guava flow cytometers (Luminex Corp), and following dilution 250,000 peripheral blood mononuclear cells per well were processed using T-SPOT *Discovery* SARS-CoV-2 tests, incubated for 16-20 hours at 37 degrees. Seven wells were used for each sample- 1. Nil Control to identify non-specific cell activation, Panels 2-6 SARS-CoV-2 specific antigen panels and Panel 7- Positive Control: Mitogen solution containing phytohaemagglutinin (PHA) to confirm PBMC functionality (Table S3). A second antibody, conjugated to alkaline phosphatase and directed to a different epitope on the cytokine molecule, is added and binds to the cytokine captured on the membrane surface. Any unbound conjugate is removed by washing. A soluble substrate is added to each well; this is cleaved by bound enzyme to form a spot of insoluble precipitate at the site of the reaction. Spots were read using automated cell counting equipment (CTL, Immunospot). The protocol followed the manufacturer's instructions exactly.

A sample was considered indeterminate if a) the Nil Control spot count was in excess of 6 spots per 250,000 PBMCs or b) the Positive Control well containing phytohemagglutinin (PHA) had less than 20 spots.

#### Masking

None of the individuals who ran the laboratory immunoassays, either serological or T-SPOT, had access to any information about the samples. The only exception is with the 154 Stream C samples. Stream C recruited after Streams A and B, and only contained PCR positive individuals. Laboratory staff knew this prior to sample receipt. All laboratory immunoassays were completed prior to any data analysis. Participants were made aware of their EUROIMMUN serological results approximately one month after their clinic visit, with a warning this was not indicative of protection from disease. Participants were not informed of their Roche or T-SPOT results. We derived the T-SPOT cutoff value, separating individuals with higher vs. lower SARS-CoV-2 responsive T cell numbers, based on separation of individuals with known SARS-CoV-2 infection vs. a low risk cohort; this activity preceded analysis of follow-up data.

#### Endpoints used on follow-up

The endpoint was having a SARS-CoV-2 RT-PCR positive test at least 12 days after recruitment.

For participants symptom driven testing was available without charge through state and employer routes. National criteria operate for obtaining such tests; these require at least one of a persistent cough, fever, or a change of taste or smell[1]. Asymptomatic screening of contacts operated in some workplaces from 6 November 2020, in response to outbreaks. Nationally, SARS-CoV-2 RT-PCR test results are gathered by Public Health England (PHE)'s Second Generation Surveillance System (SGSS) with laboratory reporting by this route is required by law. Participants consented to national data systems being searched for their records.

Participants were matched using National Health Service (NHS) number (a unique identifier used by the NHS to identify all registered individuals). For 87% of participants, we were able to look up NHS numbers, which are not widely known by UK residents, by Demographics Batch Searches against NHS data systems. In all cases, we additionally performed searches based on surname, forename and data of birth to maximise ascertainment of SARS-CoV-2 RT-PCR results.

We performed additional checks on individuals with a positive endpoint as follows:

1. All positive results were reviewed manually and correct matching assured using geographic or and any other identifiers unused in the initial match.
2. Where available, the contemporaneous recorded symptom noted by the participant was checked. A proportion of volunteers agreed on recruitment to perform weekly symptom diaries. This was optional, and volunteers could opt out.

---

[1] https://www.nhs.uk/conditions/coronavirus-covid-19/testing-and-tracing/get-a-test-to-check-if-you-have-coronavirus/ accessed 10 October 2020

3. All participants with positive tests were sent an optional post-positive test questionnaire. This asked about symptoms and circumstances around the test.

For survival analysis, we started follow-up 12 days after the recruitment clinic visit, to avoid inclusion of individuals who may have been incubating infection at the clinic visit. This resulted in the exclusion of one individual only, who developed disease 5 days after recruitment. For the logistic regression analysis, we started follow-up on 8 July 2020, which is 12 days after the last participant was recruited.

Various national surveillance projects (such as those run by the Office for National Statistics[2], and the REACT1 study[3]), do offer tests to UK citizens. We excluded one individual with chronic post-COVID-19 symptoms who took such a test without any change in their symptoms. The individual was notified of the test result, which was positive, by the national surveillance project; the participant contacted the EDSAB-HOME study team to discuss this result as they (and we, subsequently) were unclear as to its interpretation. It may reflect a false positive test.

In late Sept-2020, a second wave of infection was beginning in England; for detail, see: https://coronavirus.data.gov.uk/ and Figure S1.

#### Statistics

##### Objective 1: Describe baseline immunological responses to the SARS-CoV-2 virus

To explore and depict individuals' multi-dimensional immunological results (T-SPOT counts to six pools of peptides; immunoassay results), we performed hierarchical clustering, displaying results as heatmaps. In doing so, we transformed T-SPOT responses generating $\log_{10}$ (number of cells per 250,000 + 1), and transformed serological indexes (the raw value reported by the immunoassay) as $\log_{10}$ (immunoassay signal). This data was then clustered based on Minkowski distances using hierarchical clustering using Ward's method (4). Data were depicted using the R *hclust* function. In some analyses, we report the sum of T-SPOT responses to groups of wells, subtracting the background, see Supplementary Material Table S3. Comparing T-SPOT responses in two groups of

[2] https://www.ons.gov.uk/peoplepopulationandcommunity/healthandsocialcare/conditionsanddiseases/methodologies/covid19infectionsurveypilotmethodsandfurtherinformation accessed 10 Sept 2020

[3] https://www.imperial.ac.uk/medicine/research-and-impact/groups/react-study/ accessed 10 Sept 2020

individuals was done using Wilcoxon Ranked-sign tests, and more than two groups used Kruskal-Wallis tests.

To depict associations between immunological and clinical metadata (see Table S2), we computed Spearman correlation coefficients ρ, regarding ρ different from 0 if the associated *p* value was less than 0.01, adjusted for multiple comparisons using Bonferroni's method, depicting correlations as heatmaps.

To analyse the T-SPOT result of individuals, we computed the sum of T-SPOT responses to groups of panels, subtracting the background (Table S3). T-SPOT values optimally separating 154 individuals with high vs. 1087 individuals at low risk of past infection were computed (see Supp. Methods Fig. S6) using Youden's method (37). The high risk individuals had had previous positive RT-PCR tests. The low risk individuals reported no personal or household symptoms compatible with COVID-19 since January 2020, had had no positive RT-PCR tests, and had negative anti-Spike and anti-Nucleoprotein immunoassays using stringent cutoffs (index less than 0.35 for both assays).

#### Objective 2: Measure associations between baseline immunological responses and subsequent COVID-19

To determine whether T-SPOT results or anti-Spike IgG concentrations (immunological predictors) were associated with protection from COVID-19, we started follow-up 12 days after the last recruitment (i.e. 8 July 2020), so subjects were followed for the same time. We did this to avoid inclusion of individuals who may have developed immune responses prior to symptom recognition, choosing the 12 day cutoff because only about 2.5% of cases have incubation periods longer than 11.5 days(38). Two endpoints were excluded on these grounds (see also Supplementary Materials).

We first quantified univariable associations between a number of baseline factors, including categorised antibody index (Table S4) and T-SPOT response. We similarly divided and analysed T-SPOT SNM responses. To quantify effect sizes, we used univariable and multivariable Poisson regression models including age (as a continuous variable), gender, high risk occupation (defined as being medical or nursing hospital staff), ethnicity (white vs. non-white) as well as (for time-to-event analyses) the average incidence (individuals testing positive in the last 7 days) in the NHS region of residence of the participant included as a time-dependent covariate. We also included T-SPOT response and EuroImmun anti-Spike IgG assay result. In some analyses, we used categorisations of these (as above), and in others we modelled these as continuous variables. When modelling EuroImmun antibody indexes as continuous variables, we recoded values of over 10 as 10, as values over 10 indicate assay saturation. We then subtracted 0.35 from the variables, recoding negative values as zero, because we regard values less than 0.35 as representing technical assay background, with which disease risk is not expected to alter (see Results). We modelled square root transformed EuroImmun anti-Spike IgG index and square root transformed T-SPOT SNM responses, after selecting suitable transformations from linear, square root, log transform, 3 and 5 knot cubic splines using visual linearity and homoscedasticity assessment (by assessment of residuals), as well as Bayesian information criteria (BIC). In sensitivity analyses, we also used log transformations.

Analyses used R 4.0.2 or (for Poisson regression) Stata 16 for Windows.

#### Sample size calculations

Study size was aimed to achieve narrow confidence intervals around the performance of antibody detection tests, as described in the trial protocol (ISRCTN56609224).

#### Tables

##### Table S1: Reported Seroconversion rates in acute SARS-CoV-2 infection

ND, not determined.

| Reference | Cohort | Seroconversion estimate: Roche SARS-CoV-2 N protein immunoassay (95% CI) | Seroconversion estimate: EUROIMMUN SARS-CoV-2 S1 protein immunoassay (95% CI) | Seroconversion estimate: other assays |
|---|---|---|---|---|
| (2) | 536 positive samples from unique adult individuals with laboratory-confirmed SARS-CoV-2 infection at ≥20 days post-symptom onset. Predominantly hospitalised. | 97.2% (95% CI 95.4 to 98.4) | - | ND |
| (1) | 268 key workers with laboratory-confirmed SARS-CoV-2 infection median 63 days post-symptom onset. Predominantly community (10% hospitalised) | 96.6% (95% CI 93.7% to 98.2%) | EuroImmun: 93.3% (95% CI 89.6% to 95.7%) | ND |
| (5) | 285 hospitalised COVID-19 cases | ND | ND | 100% > 20 days after infection |
| (6) | 34 cases, 30 PCR confirmed, 32/34 were mild | ND | ND | Measured responses against RBD (part of S1). 100%, but decline noted |
| (7) | 65 hospitalised COVID-19 cases | ND | ND | 100% > 20 days after infection, but decline noted |
| (8) | 168 hospitalised COVID-19 cases | ND | ND | 100%; avidity maturation noted |
| (9) | Systematic review, n=8,256. Cases mostly hospitalised | ND | ND | 96% (90-98%) for combined IgG/IgM at d 30 + |

#### Table S2: EDSAB-HOME Demographics and exposure characteristics

| | Stream A: Police and Fire | Stream B: Healthcare workers (HCW) | Stream C: HCW Previously Positive (HCW-PP) | Total |
|---|---|---|---|---|
| **Age (years)** | | | | |
| 18 – 25 | 37 (3.2%) | 89 (5.8%) | 13 (8.4%) | 139 (4.9%) |
| 25 – 40 | 420 (36.6%) | 586 (37.9%) | 62 (40.3%) | 1068 (37.5%) |
| 40 – 60 | 660 (57.5%) | 747 (48.3%) | 71 (46.1%) | 1478 (51.9%) |
| 60+ | 30 (2.6%) | 124 (8.0%) | 8 (5.2%) | 162 (5.7%) |
| **Gender** | | | | |
| Female | 455 (39.7%) | 1247 (80.7%) | 126 (81.8%) | 1828 (64.2%) |
| Male | 692 (60.3%) | 299 (19.3%) | 28 (18.2%) | 1019 (35.8%) |
| **Ethnicity** | | | | |
| White | 1085 (94.6%) | 1128 (73.0%) | 137 (89.0%) | 2350 (82.5%) |
| Asian or British Asian | 33 (2.9%) | 237 (15.3%) | 11 (7.1%) | 281 (9.9%) |
| Black or Black British | 2 (0.2%) | 96 (6.2%) | 1 (0.6%) | 99 (3.5%) |
| Mixed | 21 (1.8%) | 42 (2.7%) | 3 (1.9%) | 66 (2.3%) |
| Other | 6 (0.5%) | 43 (2.8%) | 2 (1.3%) | 51 (1.8%) |
| **Occupation** | | | | |
| First responder – police | 528 (46.0%) | 0 (0.0%) | 0 (0.0%) | 528 (18.5%) |
| First responder – fire & rescue | 238 (20.7%) | 0 (0.0%) | 0 (0.0%) | 238 (8.4%) |
| First responder – other (e.g. ambulance) | 40 (3.5%) | 1 (0.1%) | 0 (0.0%) | 41 (1.4%) |
| Hospital doctor | 0 (0.0%) | 245 (15.8%) | 41 (26.6%) | 286 (10.0%) |
| Hospital nurse | 1 (0.1%) | 471 (30.5%) | 51 (33.1%) | 523 (18.4%) |
| Hospital medical other | 0 (0.0%) | 249 (16.1%) | 18 (11.7%) | 267 (9.4%) |
| Hospital non-medical | 0 (0.0%) | 158 (10.2%) | 9 (5.8%) | 167 (5.9%) |
| Hospital lab based | 0 (0.0%) | 35 (2.3%) | 1 (0.6%) | 36 (1.3%) |
| GP doctor/nurse/other | 0 (0.0%) | 20 (1.3%) | 4 (2.6%) | 24 (0.8%) |
| Community nurse | 0 (0.0%) | 36 (2.3%) | 9 (5.8%) | 45 (1.6%) |
| NHS staff other | 0 (0.0%) | 309 (20.0%) | 20 (13.0%) | 329 (11.6%) |
| Other | 340 (29.6%) | 22 (1.4%) | 1 (0.6%) | 363 (12.8%) |

| | | | | |
|---|---|---|---|---|
| **Interacted face-to-face with patients and/or general public during lockdown (post-23 March 2020)** | | | | |
| More frequently than before lockdown | 24 (2.1%) | 161 (10.4%) | 13 (8.4%) | 198 (7.0%) |
| Similar to before lockdown | 494 (43.1%) | 671 (43.4%) | 91 (59.1%) | 1256 (44.1%) |
| Less frequently than before lockdown | 418 (36.4%) | 564 (36.5%) | 40 (26.0%) | 1022 (35.9%) |
| No such interactions | 39 (3.4%) | 28 (1.8%) | 2 (1.3%) | 69 (2.4%) |
| Non-response | 172 (15.0%) | 122 (7.9%) | 8 (5.2%) | 302 (10.6%) |
| **Do you think you have had previous COVID-19?** | | | | |
| Yes, I had symptoms but was not tested | 225 (19.6%) | 301 (19.5%) | 0 (0.0%) | 526 (18.5%) |
| Yes, I had symptoms but my test(s) were all negative | 41 (3.6%) | 112 (7.2%) | 0 (0.0%) | 153 (5.4%) |
| Yes, I had symptoms, and I had at least one positive test | 24 (2.1%) | 80 (5.2%) | 152 (98.7%) | 256 (9.0%) |
| Yes, I had symptoms, had a test, but it failed | 5 (0.4%) | 3 (0.2%) | 0 (0.0%) | 8 (0.3%) |
| I did not recognise that I had symptoms, but I tested positive when I was screened | 0 (0.0%) | 10 (0.6%) | 2 (1.3%) | 12 (0.4%) |
| No | 557 (48.6%) | 648 (41.9%) | 0 (0.0%) | 1205 (42.3%) |
| Unsure | 295 (25.7%) | 392 (25.4%) | 0 (0.0%) | 687 (24.1%) |
| **Length of symptoms*** | | | | |
| Less than 7 days | 115 (39.1%) | 178 (36.1%) | 30 (19.7%) | 323 (34.4%) |
| 7 -14 days | 121 (41.2%) | 185 (37.5%) | 63 (41.4%) | 369 (39.3%) |
| 14 – 21 days | 27 (9.2%) | 66 (13.4%) | 27 (17.8%) | 120 (12.8%) |
| More than 21 days | 28 (9.5%) | 58 (11.8%) | 30 (19.7%) | 116 (12.4%) |
| Do not know | 3 (1.0%) | 6 (1.2%) | 2 (1.3%) | 11 (1.2%) |
| **Unable to work (in workplace or at home) due to symptoms*** | | | | |
| Yes | 191 (65.0%) | 350 (71.0%) | 144 (94.7%) | 685 (72.9%) |
| No | 103 (35.0%) | 143 (29.0%) | 8 (5.3%) | 254 (27.1%) |
| **Went to hospital due to suspected/confirmed COVID-19*** | | | | |
| Yes | 4 (1.4%) | 19 (3.9%) | 15 (9.9%) | 901 (96.0%) |
| No | 290 (98.6%) | 474 (96.1%) | 137 (90.1%) | 38 (4.0%) |
| **Was in contact with a suspected or confirmed case in the 14-day prior to symptom onset*** | | | | |
| Yes, confirmed | 23 (7.8%) | 153 (31.0%) | 92 (60.5%) | 485 (51.7%) |
| Yes, suspected | 47 (16.0%) | 113 (22.9%) | 26 (17.1%) | 268 (28.5%) |

| | | | | |
|---|---|---|---|---|
| No/Unsure | 224 (76.2%) | 227 (46.0%) | 34 (22.4%) | 186 (19.8%) |
| **Has had a SARS-CoV-2 antibody test, and been informed of the result prior to recruitment** | | | | |
| Yes | 4 (0.3%) | 20 (1.3%) | 68 (44.2%) | 92 (3.2%) |
| No | 1143 (99.7%) | 1523 (98.5%) | 86 (55.8%) | 2752 (96.7%) |
| Did not answer | 0 (0.0%) | 3 (0.2%) | 0 (0.0%) | 3 (0.1%) |
| **Had a household member who had COVID-19 compatible symptoms** | | | | |
| Yes | 289 (25.2%) | 396 (25.6%) | 83 (53.9%) | 768 (27.0%) |
| No | 858 (74.8%) | 1150 (74.4%) | 71 (46.1%) | 2079 (73.0%) |
| **Had a household member who had SARS-CoV-2 positive nasal or throat swab** | | | | |
| Yes | 21 (1.8%) | 44 (2.8%) | 32 (20.8%) | 97 (3.4%) |
| No | 1126 (98.2%) | 1502 (97.2%) | 122 (79.2%) | 2750 (96.6%) |
| **Total** | **1147** | **1546** | **154** | **2847** |

*for only those who self-reported COVID-19 compatible symptoms. In addition, variables where the participant did not respond (NAs) amongst those who reported symptoms were not included (1 in Police and Fire and 3 in HCW). Final total was 939, split across Police and Fire (n=295), HCW (n=496) and HCW-PP (n=152).

#### Table S3: Peptide panel results reported

Details of the peptide panels in the T-SPOT *Discovery* SARS-CoV-2 test

| | Response to | Reported |
|---|---|---|
| Panel 1 | S1 Spike domain | Observed cell number per 250,000 PBMC |
| Panel 2 | S2 Spike domain | Observed cell number per 250,000 PBMC |
| Panel 3 | Nucleoprotein | Observed cell number per 250,000 PBMC |
| Panel 4 | Matrix protein | Observed cell number per 250,000 PBMC |
| Panel 5 | Structural peptides | Observed cell number per 250,000 PBMC |
| Panel 6 | Envelope | Observed cell number per 250,000 PBMC |
| Control | No peptide added | Observed cell number per 250,000 PBMC |
| SNM | S antigen, Nucleoprotein and Matrix protein | (Panel1 + Panel 2 + Panel 3 + Panel 4) – 4(Control), or zero is less than zero |
| ESTR | Envelope and structural proteins | (Panel 5 + Panel 6) – 2(Control), or zero if less than zero |

#### Table S4: Categorisation of EuroImmun anti-Spike IgG index

| EuroImmun anti-SARS-CoV-2 Spike S1 domain IgG immunoassay index | Category referred to as | Comment |
|---|---|---|
| 0 <= index < 0.35 | Seronegative | Cutoff is based on comparison with pre-pandemic sera, see results |
| 0.35 <= index < 1.1 | Weak reactivity | 1.1 cutoff corresponds to the manufacturer's recommended 'confirmed positive' cutoff |
| 1.1 <= index < 3 | Low level seropositive | About 30% of the EDSAB-HOME cohort with results > 1.1 have results in this range, see results |
| Index >=3 | High level seropositive | |

#### Table S5: Responses to different peptide panels

| | | | anti-Spike index < 1.1 n=2,264 Centiles | | | | | anti-Spike index >= 1.1 n=562 Centiles | | | | | Wilcoxon test $-\log_{10}(p)$ |
|---|---|---|---|---|---|---|---|---|---|---|---|---|---|
| | Response to | Note | 5% | 25% | 50% | 75% | 95% | 5% | 25% | 50% | 75% | 95% | |
| Panel 1 | S1 Spike domain | 1 | 0 | 0 | 1 | 2 | 6 | 1 | 3 | 7 | 14 | 39 | 170.9 |
| Panel 2 | S2 Spike domain | 1 | 0 | 0 | 2 | 3 | 11 | 1 | 4 | 6 | 12 | 33 | 129.3 |
| Panel 3 | Nucleoprotein | 1 | 0 | 0 | 1 | 3 | 8 | 1 | 6 | 12 | 23 | 59 | 200.0 |
| Panel 4 | Matrix protein | 1 | 0 | 0 | 1 | 3 | 7 | 0 | 3 | 6 | 13 | 37 | 132.6 |
| Panel 5 | Structural peptides | 1 | 0 | 0 | 1 | 2 | 4 | 0 | 0 | 1 | 3 | 7 | 12.0 |
| Panel 6 | Envelope | 1 | 0 | 0 | 1 | 2 | 6 | 0 | 0 | 1 | 2 | 6 | 0.1 |
| Cont-rol | No peptide added | 1 | 0 | 0 | 0 | 1 | 3 | 0 | 0 | 0 | 1 | 3 | 0.5 |
| SNM | S antigen, Nucleoprotein and Matrix protein | 2 | 0 | 1 | 3 | 8 | 26 | 7 | 17 | 34 | 61 | 142 | 211.5 |
| ESTR | Envelope and structural proteins | 3 | 0 | 0 | 1 | 3 | 8 | 0 | 0 | 1 | 3 | 9 | 3.5 |

Note: metric reported is

1. Observed cell number per 250,000 PBMC
2. (Panel1 + Panel 2 + Panel 3 + Panel 4) – 4(Control), or zero is less than zero
3. (Panel 5 + Panel 6) – 2(Control), or zero if less than zero

#### Table S6: Risk factors for high SNM T-SPOT results when seronegative

SNM T cell responses per 250,000 PBMC, measured by the T-SPOT® *Discovery* SARS-CoV-2 test in EDSAB-HOME participants, stratified by anti-Spike IgG serostatus as well as clinical and immunological features.

| | | | All participants (n=2,826) | | | | | | | EuroImmun anti-Spike IgG index < 1.1 (n=2,264) | | | | | | | Seronegative participants: EuroImmun anti-Spike IgG index < 0.35 (n=2,172) | | | | | | | EuroImmun anti-Spike IgG positive (index >= 1.1) (n=562) | | | | | | |
|---|---|---|---|---|---|---|---|---|---|---|---|---|---|---|---|---|---|---|---|---|---|---|---|---|---|---|---|---|---|---|
| | | | T-SPOT SNM response centiles | | | | | | Kruskal-Wallis | T-SPOT SNM response centiles | | | | | | Kruskal-Wallis | T-SPOT SNM response centiles | | | | | | Kruskal-Wallis | T-SPOT SNM response centiles | | | | | | Kruskal-Wallis |
| | SARS-CoV-2 risk factor | Category | n | 5% | 25% | Median | 75% | 95% | $-\log_{10}(p)$ | n | 5% | 25% | Median | 75% | 95% | $-\log_{10}(p)$ | n | 5% | 25% | Median | 75% | 95% | $-\log_{10}(p)$ | n | 5% | 25% | Median | 75% | 95% | $-\log_{10}(p)$ |
| Demographics | Self-reported Ethnicity | Asian or Asian British | 280 | 0 | 1 | 5 | 16 | 65 | 0.09 | 210 | 0 | 0 | 3 | 6.75 | 22.55 | 1.26 | 201 | 0 | 0 | 3 | 6 | 20 | 1.27 | 70 | 5.45 | 16 | 32.5 | 56.25 | 215.5 | 0.03 |
| | | Black or Black British | 99 | 0 | 2 | 6 | 18 | 69 | - | 65 | 0 | 0 | 3 | 7 | 15.8 | - | 62 | 0 | 0 | 3 | 7.75 | 15.95 | | 34 | 6.95 | 15.25 | 29.5 | 61.5 | 124.05 | |
| | | Mixed | 66 | 0 | 2 | 6 | 18 | 62 | - | 51 | 0 | 1 | 4 | 8 | 22.5 | - | 49 | 0 | 1 | 4 | 7 | 13.8 | | 15 | 9.2 | 21.5 | 37 | 56.5 | 109.3 | |
| | | Other | 50 | 0 | 1 | 4.5 | 14.75 | 64 | - | 39 | 0 | 1 | 3 | 8 | 28.2 | - | 38 | 0 | 1 | 3 | 8.5 | 28.3 | | 11 | 8 | 18 | 33 | 61.5 | 108.5 | |
| | | White | 2331 | 0 | 1 | 5 | 14 | 66 | - | 1899 | 0 | 1 | 4 | 8 | 27 | - | 1822 | 0 | 1 | 3 | 8 | 23.95 | | 432 | 7 | 18 | 34 | 62 | 139 | |
| | Occupation | Community nurse/other | 45 | 0 | 1 | 5 | 13 | 111 | 10.90 | 37 | 0 | 0 | 3 | 9 | 31.4 | 0.46 | 36 | 0 | 0 | 3 | 7.5 | 20.25 | 0.42 | 8 | 12.2 | 35.75 | 58 | 124.5 | 204.3 | 0.74 |
| | | First responder - Fire & Rescue | 236 | 0 | 1 | 3 | 8 | 41 | - | 224 | 0 | 1 | 3 | 7 | 25.85 | - | 216 | 0 | 1 | 3 | 7 | 24.25 | | 12 | 3.85 | 18.5 | 44 | 68.75 | 122.25 | |
| | | First responder - other | 41 | 0 | 2 | 5 | 13 | 42 | - | 37 | 0 | 2 | 4 | 10 | 29 | - | 36 | 0 | 1.75 | 4 | 10 | 22.25 | | 4 | 15.3 | 16.5 | 78.5 | 143.75 | 152.75 | |
| | | First responder - Police | 522 | 0 | 1 | 4 | 11 | 43 | - | 462 | 0 | 1 | 4 | 9 | 25 | - | 448 | 0 | 1 | 4 | 9 | 25 | | 60 | 4.9 | 15.75 | 29 | 48.5 | 92.35 | |
| | | GP doctor/nurse/other | 24 | 0 | 1.75 | 4.5 | 19 | 126 | - | 17 | 0 | 0 | 4 | 5 | 16.2 | - | 17 | 0 | 0 | 4 | 5 | 16.2 | | 7 | 18 | 40.5 | 62 | 110 | 245 | |
| | | Hospital Doctor | 286 | 0 | 2.25 | 7.5 | 29 | 89 | - | 188 | 0 | 1 | 4 | 8 | 37.95 | - | 173 | 0 | 0 | 3 | 7 | 25.8 | | 98 | 8 | 19 | 32 | 59.25 | 221.35 | |
| | | Hospital Lab Based | 35 | 0 | 2 | 6 | 15.5 | 71 | - | 26 | 0 | 1.25 | 3.5 | 8 | 17.75 | - | 24 | 0 | 1.75 | 3.5 | 8.5 | 17.85 | | 9 | 3.4 | 7 | 37 | 62 | 98.6 | |
| | | Hospital Medical Other (Clinical Academic, Clinical Hosp) | 261 | 0 | 2 | 6 | 16 | 65 | - | 204 | 0 | 1 | 4 | 9 | 27.55 | - | 194 | 0 | 1 | 4 | 8 | 19.35 | | 57 | 8.8 | 16 | 33 | 55 | 99.4 | |
| | | Hospital Non-Medical (Maintenance, Catering, Portering) | 166 | 0 | 1 | 4 | 17 | 71 | - | 133 | 0 | 0 | 3 | 7 | 25.8 | - | 130 | 0 | 0 | 2.5 | 6.75 | 23.65 | | 33 | 9 | 23 | 45 | 67 | 139.2 | |
| | | Hospital Nurse | 520 | 0 | 2 | 7 | 21.25 | 71 | - | 365 | 0 | 1 | 3 | 8 | 29.8 | - | 346 | 0 | 1 | 3 | 7 | 25.75 | | 155 | 6 | 14.5 | 29 | 51.5 | 120.2 | |
| | | NHS staff other | 327 | 0 | 2 | 6 | 19 | 89 | - | 239 | 0 | 1 | 3 | 7.5 | 24.1 | - | 232 | 0 | 1 | 3 | 7 | 22.45 | | 88 | 6.35 | 18.75 | 38.5 | 70.5 | 161.5 | |
| | | Other | 363 | 0 | 1 | 4 | 9 | 37 | - | 332 | 0 | 1 | 3 | 6.25 | 18 | - | 320 | 0 | 1 | 3 | 6 | 17 | | 31 | 10.5 | 18.5 | 37 | 58 | 138 | |
| | Gender | female | 1816 | 0 | 2 | 5 | 16 | 71 | 1.86 | 1421 | 0 | 1 | 4 | 8 | 28 | 0.35 | 1367 | 0 | 1 | 3 | 7 | 23 | 0.84 | 395 | 6 | 17.5 | 35 | 62 | 139 | 0.06 |
| | | male | 1010 | 0 | 1 | 5 | 13 | 60 | - | 843 | 0 | 1 | 3 | 8 | 24 | - | 805 | 0 | 1 | 3 | 8 | 22.8 | | 167 | 7 | 17 | 33 | 60 | 154.4 | |
| | Age, categorised | 18-25 | 136 | 0 | 3 | 7 | 15.25 | 50 | 3.64 | 96 | 0 | 1 | 5 | 10.3 | 28.25 | 6.32 | 91 | 0 | 1 | 5 | 10.5 | 28.5 | 7.56 | 40 | 4.95 | 8.75 | 19.5 | 40 | 100.5 | 4.99 |
| | | 26-40 | 1066 | 0 | 2 | 6 | 16.75 | 55 | - | 853 | 0 | 1 | 4 | 9 | 28.4 | - | 818 | 0 | 1 | 4 | 9 | 27 | | 213 | 6 | 16 | 31 | 51 | 95.4 | |
| | | 41-60 | 1466 | 0 | 1 | 4 | 13 | 77 | - | 1194 | 0 | 1 | 3 | 7 | 21 | - | 1148 | 0 | 1 | 3 | 6 | 18 | | 272 | 8 | 20 | 38 | 74 | 164.25 | |
| | | >60 | 158 | 0 | 1 | 4 | 13.75 | 86 | - | 121 | 0 | 1 | 3 | 6 | 15 | - | 115 | 0 | 1 | 2 | 5 | 13.6 | | 37 | 8.6 | 23 | 45 | 77 | 141.6 | |
| | Previous positive PCR test | No | 2558 | 0 | 1 | 4 | 11 | 45 | 103 | 2241 | 0 | 1 | 3 | 8 | 25 | 10.50 | 2166 | 0 | 1 | 3 | 7 | 23 | 0.14 | 317 | 5 | 15 | 28 | 54 | 128.4 | 4.97 |
| | | Yes | 268 | 8 | 21.8 | 40.5 | 67 | 167 | | 23 | 3.4 | 22 | 38 | 48.5 | 78.4 | - | 6 | 4 | 7.25 | 8.5 | 24.75 | 36.75 | | 245 | 8 | 21 | 42 | 69 | 183 | |
| | Missed work with COVID-19 like illness | Not applicable - was not ill | 695 | 0 | 1 | 3 | 8 | 28 | 48.60 | 671 | 0 | 1 | 3 | 7 | 23.5 | 1.27 | 651 | 0 | 1 | 3 | 7 | 22 | 0.31 | 24 | 1.3 | 12.75 | 28 | 38.25 | 80.7 | 2.72 |
| | | Yes, I was unable to work at all (either at workplace or from home) | 912 | 0 | 3 | 10 | 36.25 | 103 | - | 531 | 0 | 1 | 4 | 9 | 38 | - | 499 | 0 | 1 | 4 | 8 | 25 | | 381 | 7 | 19 | 38 | 67 | 153 | |
| | | No, but I worked from home | 135 | 0 | 2 | 6 | 15 | 58 | - | 104 | 0 | 1.75 | 4 | 7.25 | 18 | - | 98 | 0 | 2 | 4 | 7 | 18 | | 31 | 8 | 15 | 25 | 52.5 | 129.5 | |
| | | No, I carried on as usual | 1037 | 0 | 1 | 4 | 10 | 44 | - | 915 | 0 | 1 | 4 | 8 | 25.3 | - | 881 | 0 | 1 | 3 | 7 | 23 | | 122 | 7 | 14 | 29 | 45.75 | 122.8 | |
| | | I am not sure/cannot remember | 47 | 0 | 0 | 4 | 10.5 | 25 | - | 43 | 0 | 0 | 3 | 8.5 | 24.3 | - | 43 | 0 | 0 | 3 | 8.5 | 24.3 | | 4 | 0.75 | 3.75 | 14.5 | 25 | 27.4 | |
| Household | Number of people in the household | 1 | 238 | 0 | 2 | 6 | 12.75 | 68 | 0.80 | 192 | 0 | 1 | 4 | 8 | 23.45 | 1.04 | 182 | 0 | 1 | 4 | 7.75 | 17.95 | 0.12 | 46 | 8 | 18 | 35 | 62 | 135.25 | 0.35 |
| | | 2 | 820 | 0 | 1 | 5 | 16 | 67 | - | 648 | 0 | 1 | 3 | 8 | 25 | - | 629 | 0 | 1 | 3 | 8 | 24 | | 172 | 6 | 17 | 37 | 60.25 | 130.8 | |
| | | 3 | 670 | 0 | 2 | 5 | 14 | 69 | - | 543 | 0 | 1 | 4 | 8 | 29.9 | - | 512 | 0 | 1 | 3 | 7 | 24.45 | | 127 | 7.3 | 21 | 33 | 67.5 | 159 | |
| | | 4 | 786 | 0 | 1 | 5 | 14 | 62 | - | 640 | 0 | 0 | 3 | 8 | 26 | - | 617 | 0 | 0 | 3 | 7 | 22.2 | | 146 | 6.25 | 16.25 | 36 | 59.75 | 126.75 | |
| | | 5 | 230 | 0 | 2 | 5 | 15 | 56 | - | 180 | 0 | 2 | 4 | 7 | 27.15 | - | 172 | 0 | 2 | 4 | 7 | 22.45 | | 50 | 6.45 | 15 | 28 | 40.75 | 123.3 | |
| | | six or more | 82 | 0 | 2.25 | 7 | 15 | 83 | - | 61 | 0 | 1 | 4 | 8 | 12 | - | 60 | 0 | 1 | 4 | 8 | 12.5 | | 21 | 8 | 16 | 39 | 71 | 194 | |
| | Number of children in the household | 0 | 1534 | 0 | 1 | 5 | 16 | 73 | 0.93 | 1204 | 0 | 1 | 3 | 8 | 26 | 0.98 | 1153 | 0 | 1 | 3 | 8 | 23 | 0.19 | 330 | 6 | 18 | 35 | 62 | 141.55 | 0.13 |
| | | 1 | 523 | 0 | 2 | 5 | 13 | 57 | - | 435 | 0 | 1 | 4 | 8 | 25.3 | - | 416 | 0 | 1 | 4 | 8 | 22 | | 88 | 8 | 15 | 31 | 57.5 | 138.65 | |
| | | 2 | 600 | 0 | 1 | 4 | 14 | 62 | - | 492 | 0 | 0 | 3 | 7 | 26 | - | 475 | 0 | 0 | 3 | 7 | 23 | | 108 | 6.35 | 17.75 | 33.5 | 62 | 178.35 | |
| | | 3 | 134 | 0 | 2.25 | 6 | 12 | 59 | - | 106 | 0 | 1.25 | 4 | 8 | 28.75 | - | 101 | 0 | 1 | 4 | 7 | 17 | | 28 | 8 | 15 | 28 | 46.5 | 112.65 | |
| | | 4 | 30 | 0 | 3 | 6 | 15.75 | 75 | - | 22 | 0 | 1 | 4 | 7.75 | 21.45 | - | 22 | 0 | 1 | 4 | 7.75 | 21.45 | | 8 | 15 | 15.75 | 36.5 | 65.75 | 157.25 | |
| | | 5 or more | 5 | 2.2 | 3 | 3 | 7 | 24 | - | 5 | 2.2 | 3 | 3 | 7 | 23.8 | - | 5 | 2.2 | 3 | 3 | 7 | 23.8 | | 0 | | | | | | |
| | Household COVID-19 disease beliefs | Yes, they had symptoms but they were not te | 568 | 0 | 2 | 6.5 | 22 | 78 | 16.30 | 397 | 0 | 1 | 3 | 7 | 28 | 0.20 | 376 | 0 | 1 | 3 | 7 | 21.25 | 0.73 | 171 | 6 | 18 | 35 | 58.5 | 160 | 0.57 |
| | | Yes, they had symptoms but their test(s) wer | 88 | 0 | 2 | 5 | 17.25 | 71 | - | 72 | 0 | 1 | 4 | 8 | 46.1 | | 68 | 0 | 1 | 3.5 | 7 | 19.3 | | 16 | 8.25 | 20.5 | 39 | 60.25 | 110.25 | |
| | | Yes, they had symptoms and they had at leas | 85 | 1 | 8 | 26 | 53 | 135 | - | 28 | 0 | 2 | 4.5 | 9.25 | 33.65 | | 24 | 0.15 | 2 | 3 | 8 | 24.9 | | 57 | 12.2 | 21 | 42 | 71 | 213.8 | |
| | | Yes, they had symptoms and had a test, but i | 6 | 3.5 | 5 | 8 | 62 | 114 | - | 3 | 3.2 | 4 | 5 | 42 | 71.6 | | 2 | 3.1 | 3.5 | 4 | 4.5 | 4.9 | | 3 | 5.6 | 8 | 11 | 68.5 | 114.5 | |
| | | Yes, they did not recognise that they had any | 11 | 1 | 4.5 | 8 | 18.5 | 38 | - | 5 | 0.4 | 2 | 3 | 6 | 9.2 | | 5 | 0.4 | 2 | 3 | 6 | 9.2 | | 6 | 7.25 | 10.5 | 18.5 | 25.75 | 43 | |
| | | No | 1685 | 0 | 1 | 4 | 11 | 51 | - | 1446 | 0 | 1 | 3 | 8 | 25 | | 1394 | 0 | 1 | 3 | 8 | 23 | | 239 | 6 | 16 | 33 | 61 | 130.3 | |
| | | I am not sure / would not know | 383 | 0 | 1 | 5 | 15 | 69 | - | 313 | 0 | 1 | 4 | 9 | 29 | | 303 | 0 | 1 | 4 | 9 | 24.8 | | 70 | 9.45 | 15 | 33 | 68 | 143.75 | |
| Personal symptoms | Believes has had COVID-19 | No | 1893 | 0 | 1 | 4 | 9 | 33 | 52.80 | 1764 | 0 | 1 | 3 | 8 | 25 | 0.08 | 1711 | 0 | 1 | 3 | 8 | 23 | 0.08 | 129 | 3 | 13 | 28 | 44 | 113 | 3.53 |
| | | Yes | 933 | 0 | 3 | 11 | 38 | 110 | - | 500 | 0 | 1 | 4 | 7.25 | 38 | | 461 | 0 | 1 | 3 | 7 | 21 | | 433 | 8 | 18 | 37 | 65 | 155 | |
| | Cough | No | 2021 | 0 | 1 | 5 | 13 | 54 | 7.13 | 1709 | 0 | 1 | 4 | 8 | 26 | 0.19 | 1646 | 0 | 1 | 3 | 8 | 23 | 0.51 | 312 | 7 | 16 | 32.5 | 58 | 133 | 0.64 |
| | | Yes | 805 | 0 | 2 | 7 | 22 | 84 | - | 555 | 0 | 1 | 3 | 8 | 30 | | 526 | 0 | 1 | 3 | 7 | 22 | | 250 | 6.45 | 18 | 37 | 64.5 | 154.1 | |
| | Fever | No | 2086 | 0 | 1 | 4 | 11 | 49 | 26.90 | 1834 | 0 | 1 | 3 | 8 | 26 | 0.22 | 1766 | 0 | 1 | 3 | 8 | 23 | 0.95 | 252 | 6 | 16 | 31.5 | 58 | 130.25 | 0.74 |
| | | Yes | 740 | 0 | 3 | 9 | 32.25 | 102 | - | 430 | 0 | 1 | 3.5 | 8 | 25.55 | | 406 | 0 | 1 | 3 | 7 | 21 | | 310 | 7 | 17.25 | 36 | 62.75 | 163.65 | |
| | Short of breath | No | 2166 | 0 | 1 | 4 | 12 | 53 | 13.10 | 1829 | 0 | 1 | 3 | 8 | 26 | 0.42 | 1761 | 0 | 1 | 3 | 8 | 23 | 0.42 | 337 | 6 | 15 | 31 | 56 | 128.4 | 3.07 |
| | | Yes | 660 | 0 | 3 | 7.5 | 29 | 99 | - | 435 | 0 | 1 | 4 | 8 | 30.6 | | 411 | 0 | 1 | 4 | 7 | 21 | | 225 | 8 | 20 | 41 | 69 | 166 | |
| | Sore throat | No | 1713 | 0 | 1 | 5 | 14 | 62 | 0.29 | 435 | 0 | 1 | 4 | 8 | 30.6 | 0.03 | 1325 | 0 | 1 | 3 | 8 | 22 | 0.76 | 327 | 7 | 18 | 34 | 60.5 | 149.6 | 0.10 |
| | | Yes | 1113 | 0 | 1 | 5 | 15 | 73 | - | 1386 | 0 | 1 | 4 | 8 | 26 | - | 847 | 0 | 1 | 3 | 7 | 23 | | 235 | 6 | 15 | 33 | 62.5 | 140.3 | |
| | Runny nose | No | 2040 | 0 | 1 | 5 | 14 | 65 | 2.10 | 878 | 0 | 1 | 3 | 8 | 27.15 | 3.09 | 1566 | 0 | 1 | 3 | 7 | 21 | 0.00 | 401 | 7 | 18 | 35 | 62 | 151 | 0.56 |
| | | Yes | 786 | 0 | 2 | 6 | 15 | 69 | - | 1639 | 0 | 1 | 3 | 8 | 25.1 | - | 606 | 0 | 1 | 4 | 8 | 25 | | 161 | 7 | 15 | 31 | 58 | 138 | |
| | Headache | No | 1514 | 0 | 1 | 4 | 11 | 47 | 14.10 | 625 | 0 | 1 | 4 | 8 | 29 | 1.39 | 1267 | 0 | 1 | 3 | 7 | 24 | 0.13 | 197 | 5 | 16 | 32 | 53 | 109.4 | 1.01 |
| | | Yes | 1312 | 0 | 2 | 6.5 | 20 | 89 | - | 1317 | 0 | 1 | 3 | 8 | 26 | - | 905 | 0 | 1 | 4 | 8 | 22 | | 365 | 8 | 17 | 35 | 66 | 154.6 | |
| | Muscle aches | No | 1799 | 0 | 1 | 4 | 10 | 45 | 23.40 | 947 | 0 | 1 | 4 | 8 | 27 | 1.03 | 1538 | 0 | 1 | 3 | 7 | 24 | 0.46 | 213 | 5 | 16 | 29 | 53 | 128.8 | 1.37 |
| | | Yes | 1027 | 0 | 2 | 8 | 26.5 | 96 | - | 1586 | 0 | 1 | 3 | 7 | 26 | - | 634 | 0 | 1 | 4 | 8 | 22 | | 349 | 7 | 18 | 37 | 66 | 152.6 | |
| | Abnormal sense of smell | No | 2310 | 0 | 1 | 4 | 10 | 40 | 91.80 | 678 | 0 | 1 | 4 | 9 | 27 | 2.85 | 2031 | 0 | 1 | 3 | 7 | 22 | 0.51 | 215 | 5 | 15 | 29 | 51 | 125.9 | 2.46 |
| | | Yes | 516 | 0 | 8.75 | 23 | 53 | 138 | - | 2095 | 0 | 1 | 3 | 8 | 24 | - | 141 | 0 | 1 | 4 | 9 | 30 | | 347 | 8 | 18 | 38 | 66.5 | 145.4 | |
| | Abnormal sense of taste | No | 2289 | 0 | 1 | 4 | 10 | 42 | 83.80 | 169 | 0 | 1 | 5 | 14 | 56.6 | 3.81 | 2002 | 0 | 1 | 3 | 7 | 22 | 0.23 | 225 | 5 | 15 | 29 | 51 | 127.4 | 2.36 |
| | | Yes | 537 | 0 | 7 | 22 | 49 | 137 | - | 2064 | 0 | 1 | 3 | 8 | 23 | - | 170 | 0 | 1 | 4 | 9 | 31.1 | | 337 | 7.8 | 18 | 38 | 65 | 144.4 | |
| | Fatigue | No | 1897 | 0 | 1 | 4 | 10 | 42 | 30.30 | 200 | 0 | 1 | 5 | 13 | 56.05 | 0.51 | 1628 | 0 | 1 | 3 | 8 | 22.65 | 0.95 | 211 | 5 | 15 | 28 | 47 | 96.5 | 4.73 |
| | | Yes | 929 | 0 | 3 | 9 | 33 | 111 | - | 1686 | 0 | 1 | 3 | 8 | 25 | - | 544 | 0 | 1 | 3 | 7 | 23.85 | | 351 | 8 | 19.5 | 38 | 70 | 163.5 | |
| | Diarrhoea | No | 2431 | 0 | 1 | 5 | 14 | 62 | 4.30 | 578 | 0 | 1 | 4 | 8 | 34.3 | 0.77 | 1897 | 0 | 1 | 3 | 7 | 22 | 0.14 | 454 | 6 | 16 | 33 | 59.75 | 139 | 1.64 |
| | | Yes | 395 | 0 | 2 | 7 | 24 | 92 | - | 1977 | 0 | 1 | 3 | 8 | 26 | - | 275 | 0 | 1 | 4 | 8 | 24.3 | | 108 | 11 | 21.5 | 41.5 | 71.5 | 162.8 | |
| | Nausea or Vomiting | No | 2581 | 0 | 1 | 5 | 14 | 62 | 3.08 | 287 | 0 | 1 | 4 | 8 | 38.1 | 0.57 | 2007 | 0 | 1 | 3 | 7.5 | 23 | 0.34 | 488 | 7 | 17 | 33 | 60.25 | 142.6 | 0.38 |
| | | Yes | 245 | 0 | 2 | 7 | 23 | 99 | - | 2093 | 0 | 1 | 3 | 8 | 26 | - | 165 | 0 | 1 | 4 | 8 | 21.8 | | 74 | 6 | 19.5 | 37.5 | 69 | 141.35 | |
| | Chilblains | No | 2738 | 0 | 1 | 5 | 15 | 65 | 0.96 | 171 | 0 | 1 | 4 | 8 | 25.5 | 0.23 | 2112 | 0 | 1 | 3 | 8 | 23 | 0.81 | 535 | 6 | 16 | 33 | 61 | 139 | 1.22 |
| | | Yes | 88 | 0 | 2 | 6.5 | 25.75 | 128 | - | 2203 | 0 | 1 | 3 | 8 | 26.9 | - | 60 | 0 | 1 | 3 | 7 | 21.2 | | 27 | 12.2 | 28 | 46 | 80 | 185.9 | |
| Household symptoms | Cough in household | No | 2354 | 0 | 1 | 5 | 14 | 62 | 1.95 | 1929 | 0 | 1 | 4 | 8 | 26 | 0.20 | 1857 | 0 | 1 | 3 | 8 | 23 | 0.17 | 425 | 7 | 18 | 35 | 62 | 140.6 | 0.49 |
| | | Yes | 472 | 0 | 2 | 6 | 21 | 77 | - | 335 | 0 | 1 | 3 | 7 | 31.3 | | 315 | 0 | 1 | 3 | 7 | 22 | | 137 | 6 | 16 | 31 | 57 | 146.6 | |
| | Fever in household | No | 2125 | 0 | 1 | 5 | 14 | 65 | 1.30 | 1745 | 0 | 1 | 4 | 8 | 26 | 0.47 | 1678 | 0 | 1 | 3 | 8 | 24 | 0.16 | 380 | 7 | 17.75 | 34 | 63.25 | 144.1 | 0.24 |
| | | Yes | 701 | 0 | 2 | 6 | 18 | 73 | - | 519 | 0 | 1 | 3 | 7 | 26 | - | 494 | 0 | 1 | 3 | 7 | 21 | | 182 | 6.05 | 16.25 | 33 | 56.5 | 138.45 | |
| | Short of breath in household | No | 2431 | 0 | 1 | 5 | 14 | 65 | 1.44 | 1971 | 0 | 1 | 3 | 8 | 26 | 0.22 | 1895 | 0 | 1 | 3 | 8 | 23 | 0.87 | 460 | 7 | 16.75 | 35 | 62.25 | 151.05 | 0.57 |
| | | Yes | 395 | 0 | 2 | 6 | 18.5 | 74 | - | 293 | 0 | 1 | 4 | 8 | 32.4 | - | 277 | 0 | 1 | 3 | 7 | 22 | | 102 | 6.05 | 17.25 | 31 | 51.75 | 109.85 | |
| | Sore throat in household | No | 2074 | 0 | 1 | 5 | 14 | 62 | 0.03 | 1667 | 0 | 1 | 3 | 8 | 26.7 | 0.27 | 1593 | 0 | 1 | 3 | 8 | 23 | 0.83 | 407 | 7 | 16 | 33 | 60 | 130 | 0.64 |
| | | Yes | 752 | 0 | 1 | 5 | 15 | 77 | - | 597 | 0 | 1 | 3 | 7 | 25 | - | 579 | 0 | 1 | 3 | 7 | 23 | | 155 | 6 | 18 | 37 | 69 | 170.9 | |
| | Runny nose in household | No | 2339 | 0 | 1 | 5 | 15 | 67 | 0.37 | 1869 | 0 | 1 | 3 | 8 | 27 | 1.37 | 1786 | 0 | 1 | 3 | 7 | 22 | 0.01 | 470 | 7 | 18 | 34 | 61.75 | 140.1 | 0.52 |
| | | Yes | 487 | 0 | 2 | 5 | 13 | 63 | - | 395 | 0 | 1 | 4 | 8 | 25 | - | 386 | 0 | 1 | 4 | 8 | 24.75 | | 92 | 6 | 13.75 | 31.5 | 60.25 | 175.8 | |
| | Headache in household | No | 2025 | 0 | 1 | 5 | 14 | 60 | 2.85 | 1665 | 0 | 1 | 3 | 8 | 26 | 0.03 | 1605 | 0 | 1 | 3 | 8 | 24 | 0.54 | 360 | 6 | 16 | 32.5 | 58 | 130 | 1.65 |
| | | Yes | 801 | 0 | 2 | 6 | 19 | 89 | - | 599 | 0 | 1 | 4 | 8 | 25.1 | - | 567 | 0 | 1 | 4 | 7 | 20 | | 202 | 8 | 19 | 37 | 71.75 | 164.5 | |
| | Muscle aches in household | No | 2168 | 0 | 1 | 5 | 13 | 60 | 6.53 | 1800 | 0 | 1 | 3 | 8 | 26 | 0.33 | 1736 | 0 | 1 | 3 | 8 | 23 | 0.89 | 368 | 6 | 15.75 | 32.5 | 60 | 140.3 | 1.11 |
| | | Yes | 658 | 0 | 2 | 7 | 23 | 81 | - | 464 | 0 | 1 | 4 | 8 | 28.85 | - | 436 | 0 | 1 | 4 | 7 | 22.25 | | 194 | 8 | 19 | 39 | 63.75 | 153.7 | |
| | Abnormal sense of smell in household | No | 2582 | 0 | 1 | 5 | 13 | 60 | 18.30 | 2151 | 0 | 1 | 3 | 8 | 26 | 0.37 | 2068 | 0 | 1 | 3 | 8 | 22 | 0.73 | 431 | 6 | 17 | 34 | 60 | 138.5 | 0.24 |
| | | Yes | 244 | 0 | 4 | 15 | 44 | 125 | - | 113 | 0 | 1 | 4 | 9 | 37.4 | - | 104 | 0 | 1 | 4 | 7.25 | 32.4 | | 131 | 8 | 16.5 | 36 | 65 | 203 | |
| | Abnormal sense of taste in household | No | 2583 | 0 | 1 | 5 | 13 | 61 | 14.50 | 2136 | 0 | 1 | 3 | 8 | 26 | 0.77 | 2056 | 0 | 1 | 3 | 7 | 22 | 0.48 | 447 | 6 | 17.5 | 33 | 60 | 138 | 0.44 |
| | | Yes | 243 | 0 | 4 | 12 | 39.5 | 125 | - | 128 | 0 | 1 | 4 | 9 | 37.65 | - | 116 | 0 | 1 | 4 | 8 | 33.75 | | 115 | 8 | 16 | 37 | 71 | 201.4 | |
| | Fatigue in household | No | 2264 | 0 | 1 | 5 | 13 | 62 | 3.62 | 1854 | 0 | 1 | 3 | 8 | 25 | 0.41 | 1790 | 0 | 1 | 3 | 7 | 22 | 0.99 | 410 | 7 | 16 | 33 | 61 | 142.2 | 0.33 |
| | | Yes | 562 | 0 | 2 | 7 | 22 | 79 | - | 410 | 0 | 1 | 4 | 9 | 35.1 | - | 382 | 0 | 1 | 3 | 8 | 26.9 | | 152 | 6.55 | 19 | 37 | 61.75 | 141.45 | |
| | Diarrhoea in household | No | 2604 | 0 | 1 | 5 | 15 | 65 | 0.47 | 2090 | 0 | 1 | 3 | 8 | 26 | 0.42 | 2005 | 0 | 1 | 3 | 7 | 23 | 0.25 | 514 | 7 | 18 | 34 | 61 | 139 | 0.06 |
| | | Yes | 222 | 0 | 1.25 | 6 | 14 | 87 | - | 174 | 0 | 1 | 4 | 8 | 34.7 | - | 167 | 0 | 1 | 4 | 8 | 24.4 | | 48 | 6 | 13 | 31.5 | 64.5 | 184.55 | |
| | Nausea or Vomiting in household | No | 2669 | 0 | 1 | 5 | 15 | 66 | 0.21 | 2138 | 0 | 1 | 3 | 8 | 26 | 0.30 | 2050 | 0 | 1 | 3 | 7.75 | 23 | 0.19 | 531 | 7 | 17.5 | 34 | 61.5 | 143 | 0.46 |
| | | Yes | 157 | 0 | 2 | 5 | 14 | 59 | - | 126 | 0 | 1.25 | 4 | 7.75 | 23 | - | 122 | 0 | 1.25 | 4 | 7.75 | 22.9 | | 31 | 7 | 14 | 29 | 53 | 112.5 | |
| | Chilblains in household | No | 2785 | 0 | 1 | 5 | 15 | 66 | 0.19 | 2234 | 0 | 1 | 3 | 8 | 26 | 0.25 | 2143 | 0 | 1 | 3 | 8 | 23 | 0.45 | 551 | 6.5 | 17 | 34 | 61 | 141.5 | 0.13 |
| | | Yes | 41 | 0 | 2 | 5 | 31 | 75 | - | 30 | 0 | 0.25 | 3 | 6 | 40 | - | 29 | 0 | 0 | 3 | 6 | 40 | | 11 | 10 | 20.5 | 31 | 64.5 | 136.5 | |
| | Serostatus (EuroImmun) using manufacturer's cutoff | Confirmed positive (index >=1.1) | 2264 | 0 | 1 | 3 | 8 | 26 | 211.47 | | | | | | | | | | | | | | | | | | | | | |
| | | Not confirmed positive (index < 1.1) | 562 | 7 | 17 | 34 | 61 | 142 | | | | | | | | | | | | | | | | | | | | | | |
| | Serostatus (EuroImmun) in four strata | Seronegative (index < 0.35) | 2172 | 0 | 1 | 3 | 8 | 23 | 220.30 | | | | | | | | | | | | | | | | | | | | | |
| | | Weak reactivity (0.35 <= index < 1.1) | 92 | 0 | 4 | 10 | 38 | 81 | | | | | | | | | | | | | | | | | | | | | | |
| | | Low level seropositive (1.1 <= index < 3) | 160 | 4.9 | 15 | 28.5 | 45 | 91 | | | | | | | | | | | | | | | | | | | | | | |
| | | High level seropositive (index >=3) | 402 | 7 | 18 | 36 | 67 | 155 | | | | | | | | | | | | | | | | | | | | | | |

Note:

p-values are expressed as -log10(p) values due to small numbers. Values are coloured red if they exceed a cutoff of 0.05, adjusted for having conducted 69 tests, using Bonferroni's method. This corresponds to a -log10(p) cutoff of 3.14

#### Table S7: Continuous exposure variables: primary and sensitivity analyses

Poisson regression modelling of the relationship between outcome and immunological correlates of protection. Cell values are incidence rate ratios with 95% confidence intervals. There is statistical evidence from likelihood ratio tests that models including a T cell number and antibody signal interaction term improve model fit over predictions of a model including antibodies alone (cells marked *). Correlates of protection are modelled as log transformed variables.

| | Model 1: antibodies only | | Model 2: T cells only | | Model 3: antibodies and T cells | | Model 4: antibodies, T cells and interaction | |
|---|---|---|---|---|---|---|---|---|
| | Unadjusted | With adjustment for covariates | Unadjusted | With adjustment for covariates | Unadjusted | With adjustment for covariates | Unadjusted | With adjustment for covariates |
| log(Anti-Spike IgG antibody index) | 0.57 (0.49, 0.66) | 0.54 (0.46, 0.63) | N/A | N/A | 0.58 (0.49, 0.68) | 0.55 (0.46, 0.64) | 1.10 (0.80, 1.51) | 1.06 (0.76, 1.47) |
| Log(T-SPOT response to SNM pools) | N/A | N/A | 0.77 (0.70, 0.84) | 0.75 (0.69, 0.83) | 0.97 (0.87, 1.09) | 0.98 (0.87, 1.09) | 0.49 (0.34, 0.70) | 0.48 (0.33, 0.70) |
| Interaction | N/A | N/A | N/A | N/A | N/A | N/A | 0.75 (0.65, 0.87) | 0.75 (0.64, 0.87) |
| *p*-value from a likelihood ratio test, comparing model fit vs Model 1 | N/A | N/A | N/A | N/A | 0.60 | 0.68 | 0.0002 * | 0.0002 * |

### Figures

### Figure Legends

#### Figure S1: Outbreak in the UK and its relationship to follow-up

The outbreak kinetics in the UK, and its relationship to follow-up, are shown. Data is from coronavirus.data.go.uk.

#### Figure S2: Patterns of serological responses seen

The EuroImmun anti-Spike S1 IgG index is plotted against the Roche Nucleoprotein total antibody assay ratio. The manufacturer's recommended cutoffs are shown (1.0, 1.1 respectively), as is an exploratory cutoff for the EuroImmun assay (0.3, green line).

#### Figure S3: Correlations between responses to different SARS-CoV-2 components

Spearman correlation coefficients between the S1, S2, Nucleoprotein, Matrix protein, structural proteins and Envelope proteins, as well as anti-Nucleoprotein and S1 serological responses in 2,826 individuals from streams A,B and C.

#### Figure S4: SNM T-SPOT results compared with anti-Nucleoprotein antibody

SNM (Spike, Nucleoprotein and Matrix protein) responsive T cell numbers and their relationship to anti-Nucleoprotein serological responses in 2,672 from Streams A,B. (A) distribution of anti-N (Roche) serological assay index, and its relationship to symptoms. The vertical line is at 0.8, the manufacturer's 'borderline' cutoff. (B) bivariate plots of anti-N antibody responses and SNM T-SPOT results. The horizontal line corresponds to 12 spots / 250,000 cells. (C) distribution of SNM T-SPOT results. (D) As in B, but for Envelope and Structural proteins T-SPOT results; (E) distribution of Envelope and Structural (ES) T-SPOT results. In marginal histograms (A,C,E), the number of individuals reporting symptoms is depicted in red.

#### Figure S5: SNM responsive T cell numbers in individuals with previous positive PCR tests

SNM T-SPOT results and their relationship to anti-Spike S1 (EuroImmun) serological responses in 154 individuals with past SARS-CoV-2 infection from Stream C. (A) distribution of anti-Spike S1 (EuroImmun) serological assay index, and its relationship to symptoms. The vertical line is at 0.8, the manufacturer's 'borderline' cutoff. (B) bivariate plots of anti-S1 IgG responses and SNM T-SPOT results. The horizontal line corresponds to 12 spots / 250,000 cells. (C) distribution of SNM T-SPOT results. (D) As in B, but for Envelope and Structural (ES) proteins T-SPOT results; (E) distribution of Envelope and Structural (ES) T-SPOT results. In marginal histograms (A,C,E), the number of individuals reporting symptoms is depicted in red.

#### Figure S6: Selection of cohorts at high and low risk of previous SARS-CoV-2 infection

Selection of high SARS-CoV-2 and low SARS-CoV-2 infection risk cohorts (A). (B) SNM T-SPOT results in these populations; (C) anti-S1 IgG responses in these populations; (D) bivariate plot showing anti-S1 IgG responses vs. SNM T-SPOT results. The blue box depicts individuals with elevated (more than 12 cells per 250,000 PBMC) responses who are seronegative.

#### Figure S7: ROC analysis separating low risk individuals from those with past infection

Receiver-operator curves showing separation of 1,126 individuals at low risk from historical SARS-CoV-2 infection from 154 individuals with proven infection by (A) SNM T-SPOT results; (B) Envelope and Structural protein (ES) T-SPOT results.

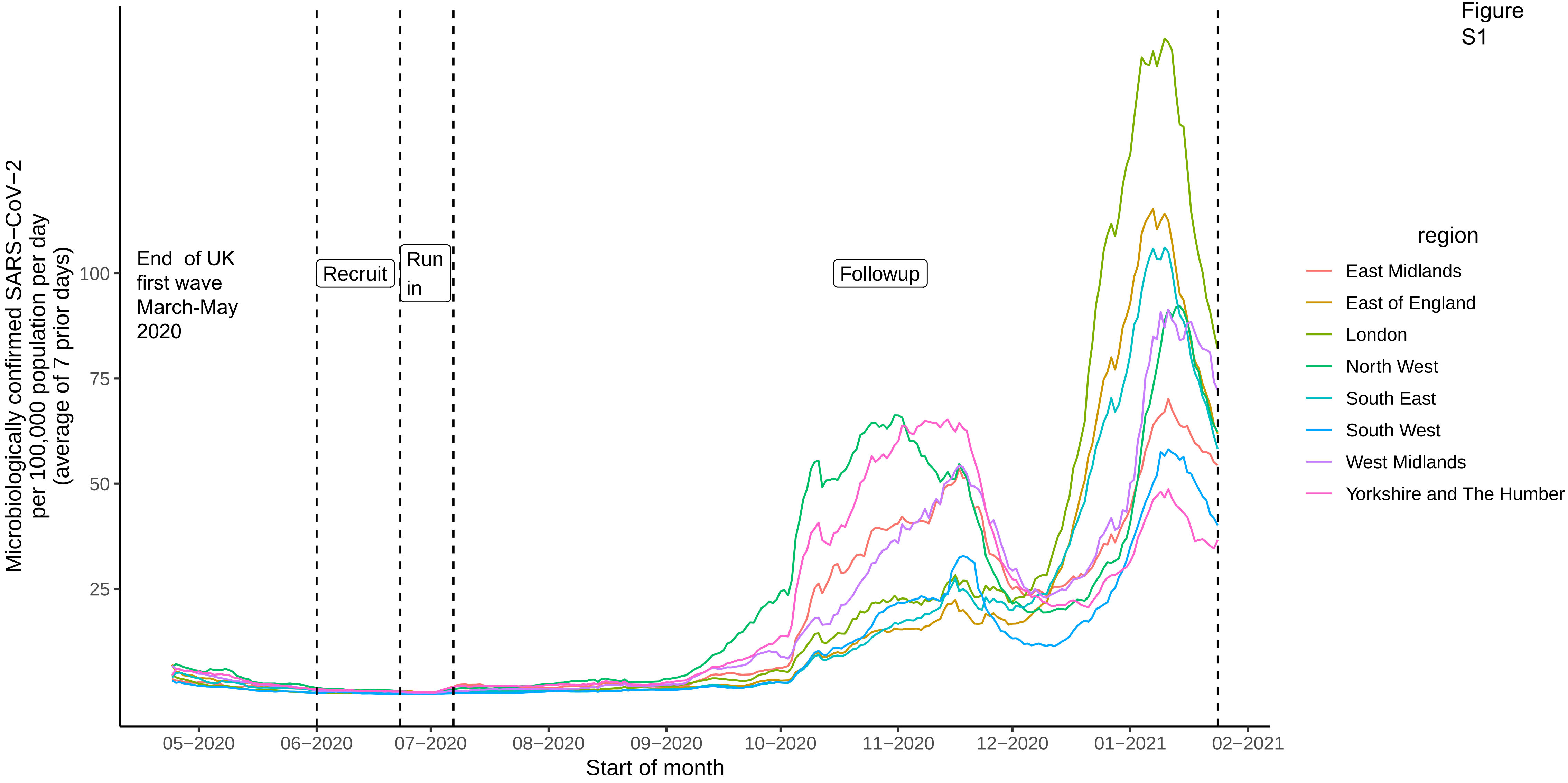
Figure S1
Microbiologically confirmed SARS−CoV−2 per 100,000 population per day (average of 7 prior days)
100
75
50
25
End of UK first wave March-May 2020
Recruit
Run in
Followup
05−2020
06−2020
07−2020
08−2020
09−2020
10−2020
11−2020
12−2020
01−2021
02−2021
Start of month
region
East Midlands
East of England
London
North West
South East
South West
West Midlands
Yorkshire and The Humber

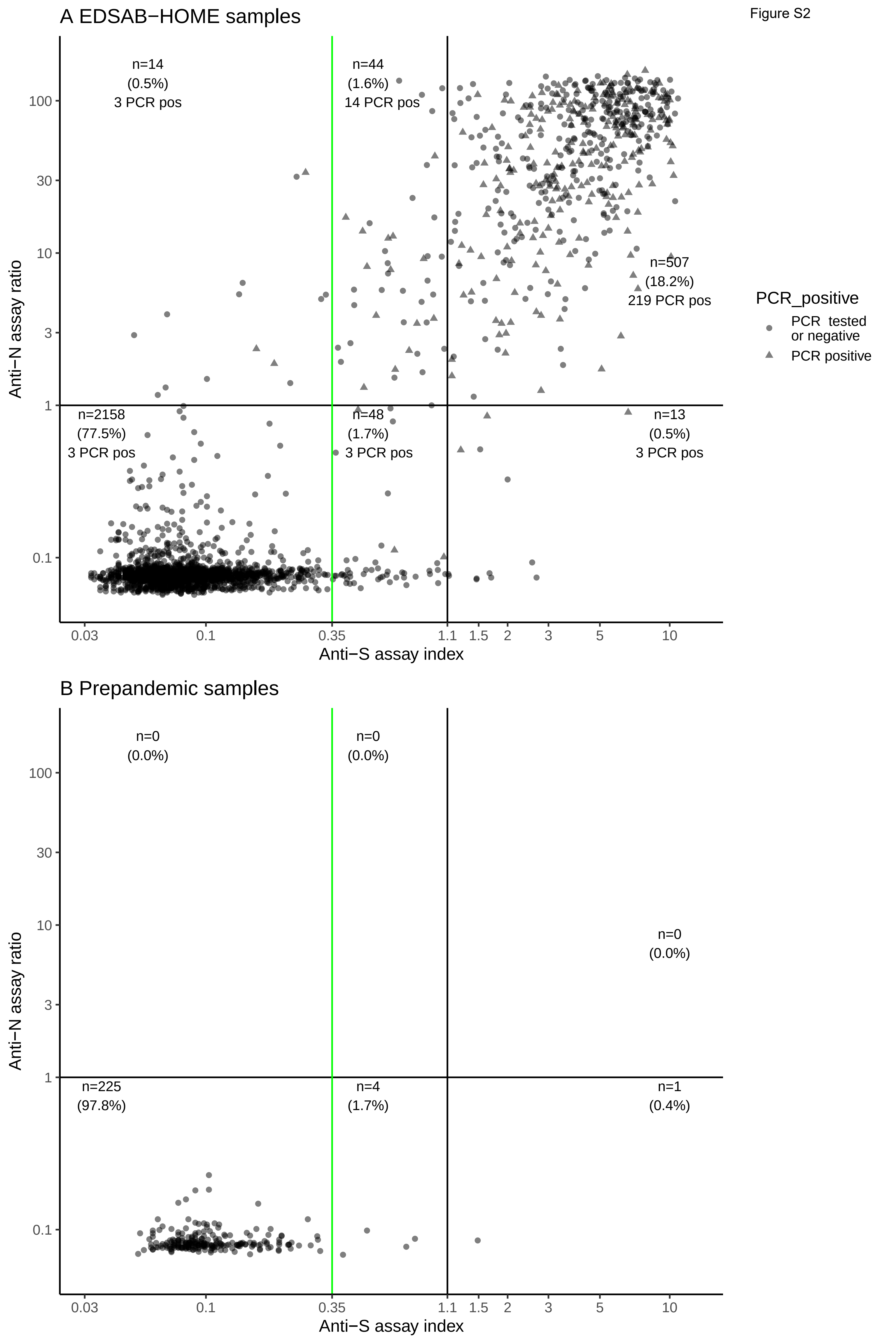
A EDSAB−HOME samples
n=14
(0.5%)
3 PCR pos
n=44
(1.6%)
14 PCR pos
n=507
(18.2%)
219 PCR pos
n=2158
(77.5%)
3 PCR pos
n=48
(1.7%)
3 PCR pos
n=13
(0.5%)
3 PCR pos
Anti−N assay ratio
Anti−S assay index
0.03
0.1
0.35
1.1
1.5
2
3
5
10
100
30
10
3
1
0.1
PCR_positive
PCR tested or negative
PCR positive
B Prepandemic samples
n=0
(0.0%)
n=0
(0.0%)
n=0
(0.0%)
n=225
(97.8%)
n=4
(1.7%)
n=1
(0.4%)
Anti−N assay ratio
Anti−S assay index

### Fig. S3

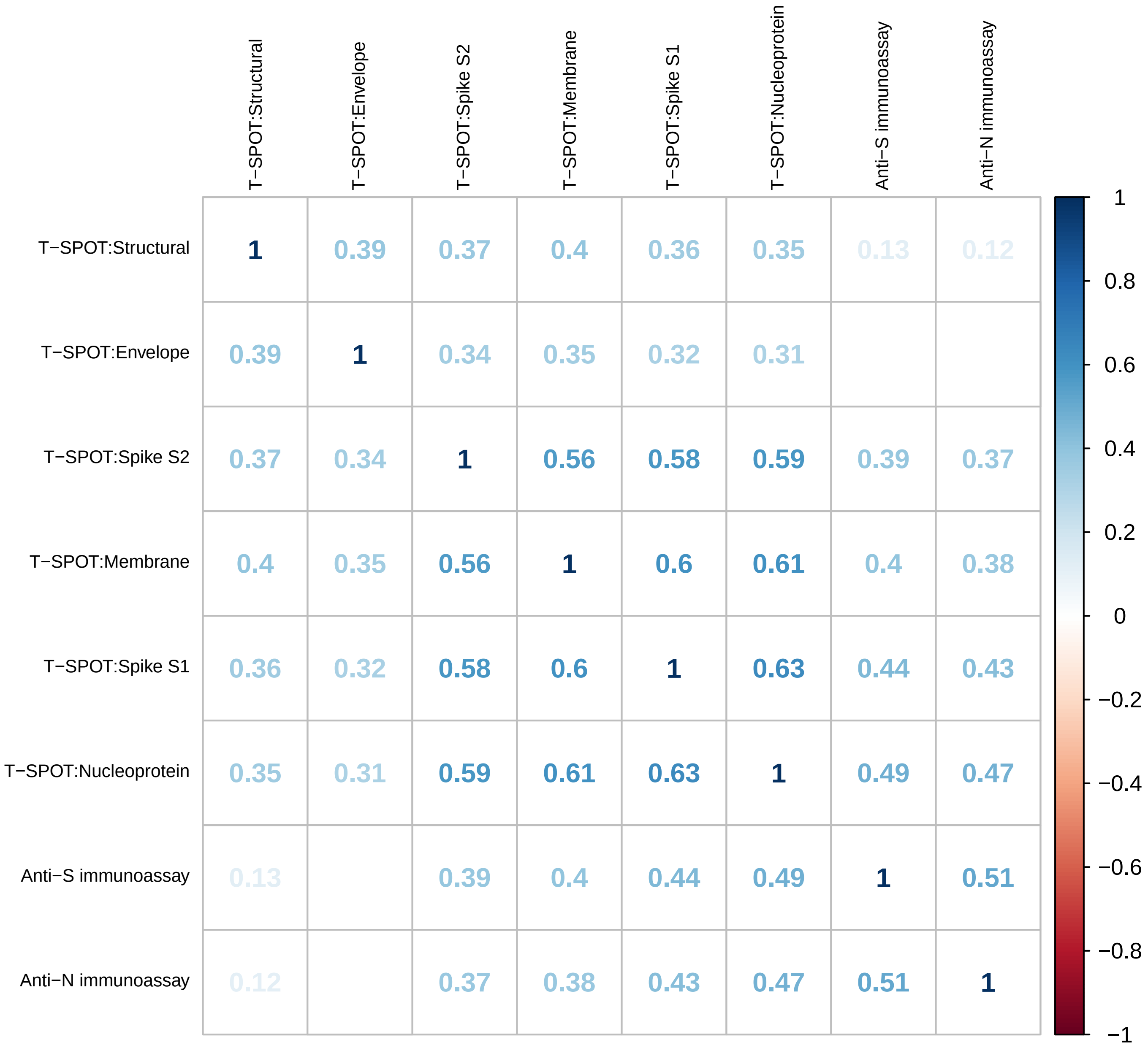

T−SPOT:Structural
T−SPOT:Envelope
T−SPOT:Spike S2
T−SPOT:Membrane
T−SPOT:Spike S1
T−SPOT:Nucleoprotein
Anti−S immunoassay
Anti−N immunoassay
1 0.39 0.37 0.4 0.36 0.35 0.13 0.12
0.39 1 0.34 0.35 0.32 0.31
0.37 0.34 1 0.56 0.58 0.59 0.39 0.37
0.4 0.35 0.56 1 0.6 0.61 0.4 0.38
0.36 0.32 0.58 0.6 1 0.63 0.44 0.43
0.35 0.31 0.59 0.61 0.63 1 0.49 0.47
0.13 0.39 0.4 0.44 0.49 1 0.51
0.12 0.37 0.38 0.43 0.47 0.51 1
1
0.8
0.6
0.4
0.2
0
−0.2
−0.4
−0.6
−0.8
−1

Figure S4

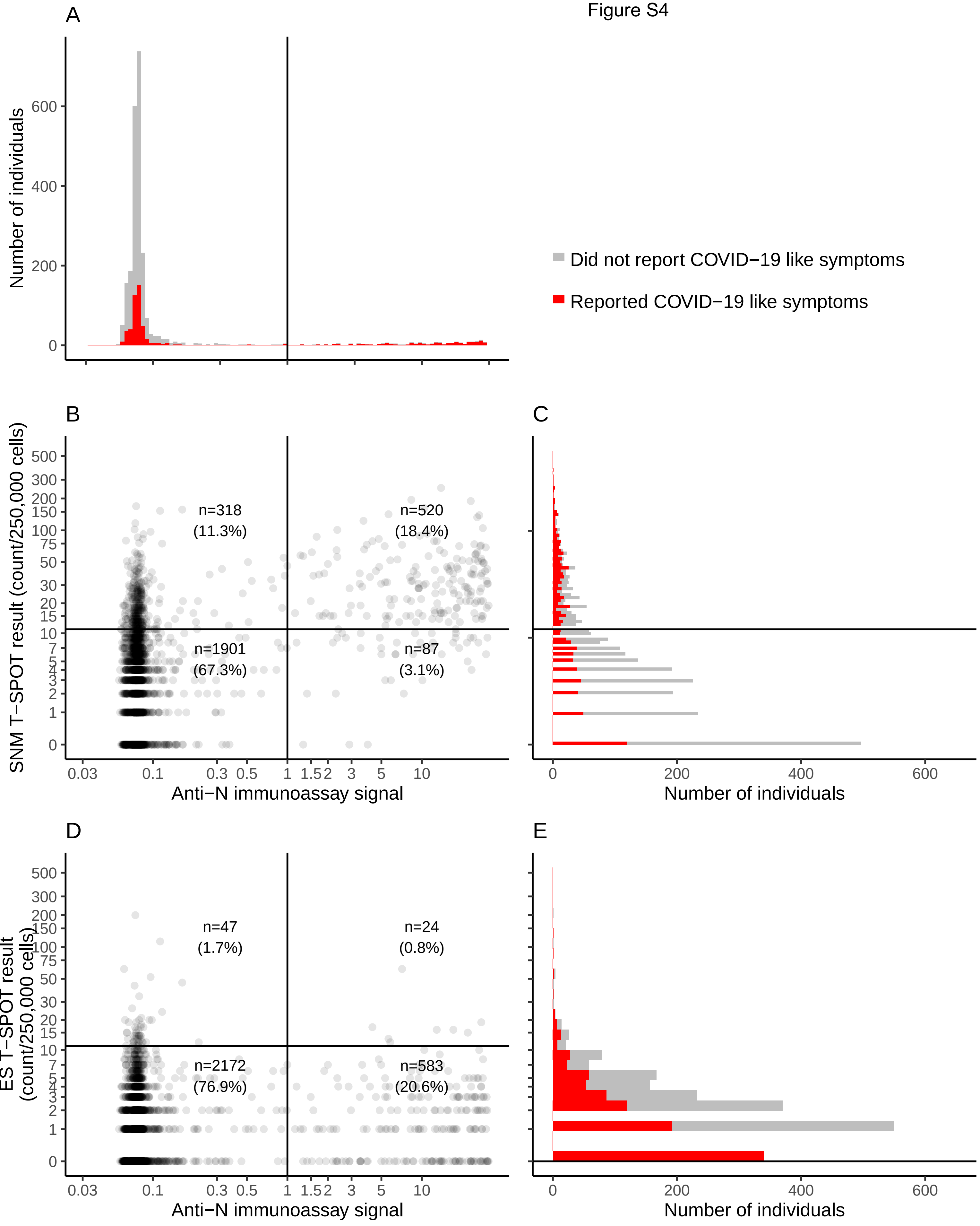
A
Number of individuals
600
400
200
0
Did not report COVID−19 like symptoms
Reported COVID−19 like symptoms
B
SNM T−SPOT result (count/250,000 cells)
n=318
(11.3%)
n=520
(18.4%)
n=1901
(67.3%)
n=87
(3.1%)
Anti−N immunoassay signal
0.03 0.1 0.3 0.5 1 1.5 2 3 5 10
C
Number of individuals
0 200 400 600
D
ES T−SPOT result (count/250,000 cells)
n=47
(1.7%)
n=24
(0.8%)
n=2172
(76.9%)
n=583
(20.6%)
Anti−N immunoassay signal
E
Number of individuals

Figure S5

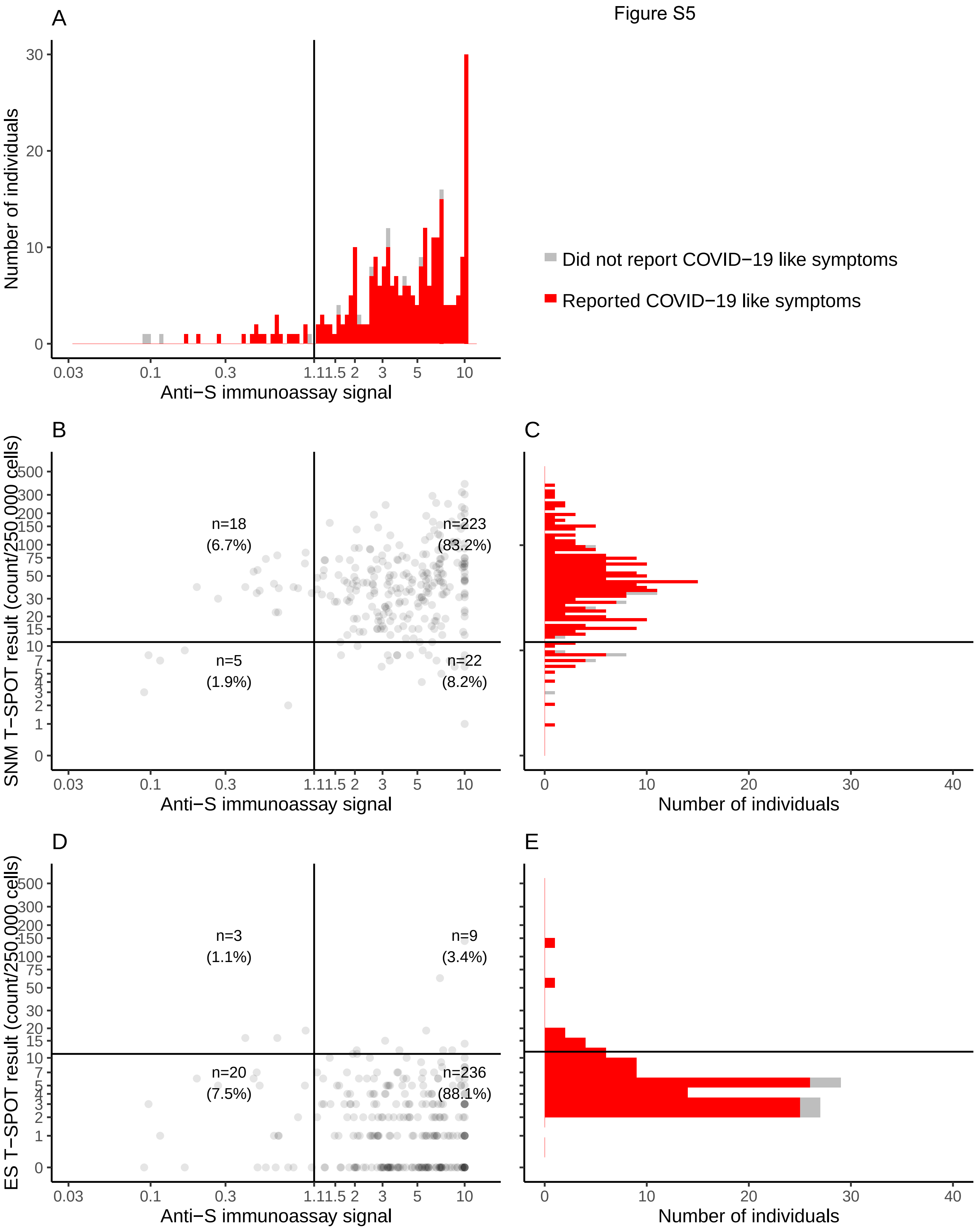
A
Number of individuals
Anti−S immunoassay signal
Did not report COVID−19 like symptoms
Reported COVID−19 like symptoms
B
SNM T−SPOT result (count/250,000 cells)
n=18
(6.7%)
n=223
(83.2%)
n=5
(1.9%)
n=22
(8.2%)
Anti−S immunoassay signal
C
Number of individuals
D
ES T−SPOT result (count/250,000 cells)
n=3
(1.1%)
n=9
(3.4%)
n=20
(7.5%)
n=236
(88.1%)
Anti−S immunoassay signal
E
Number of individuals

### Figure S6

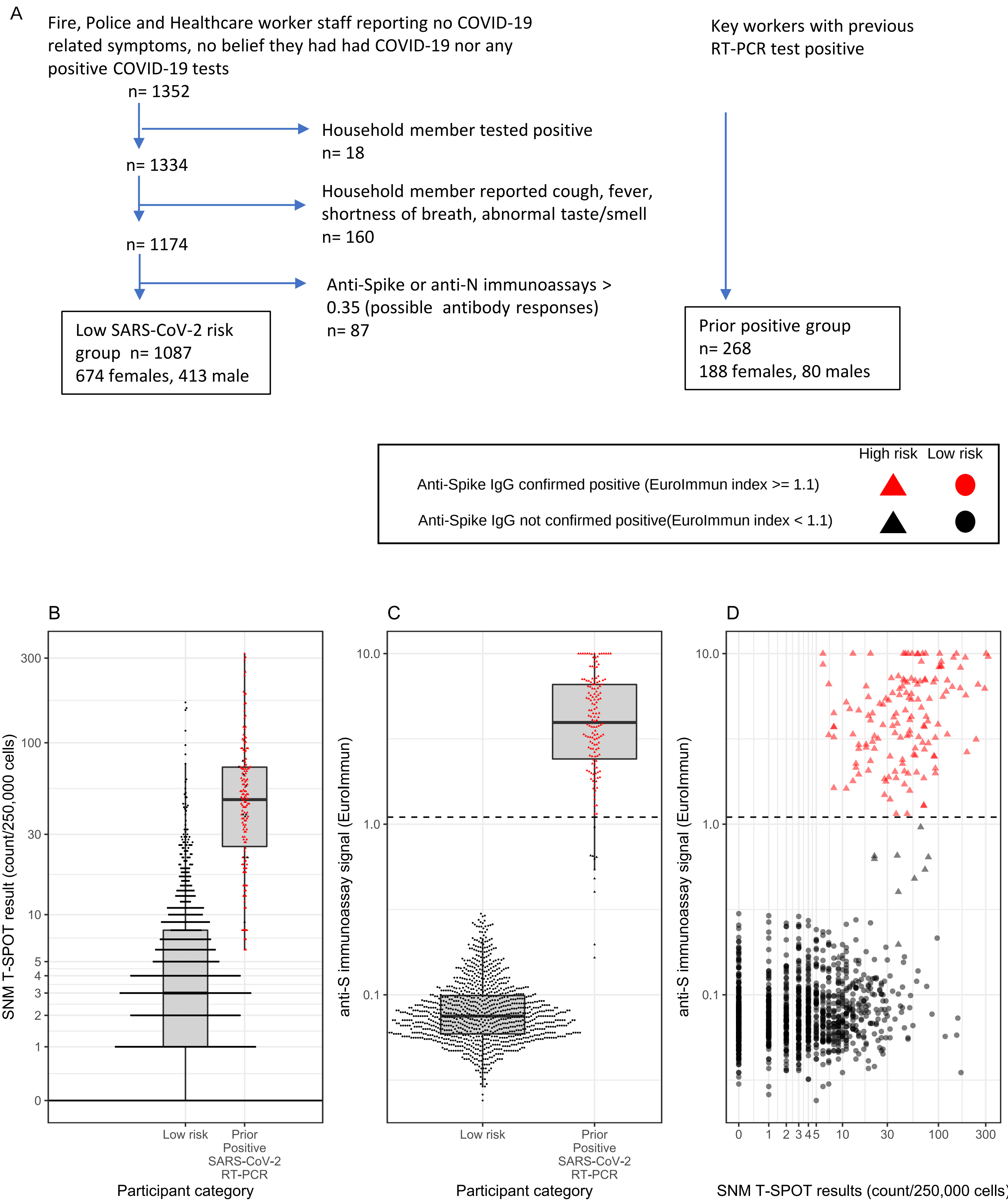
A
Fire, Police and Healthcare worker staff reporting no COVID-19 related symptoms, no belief they had had COVID-19 nor any positive COVID-19 tests
n= 1352
Household member tested positive
n= 18
n= 1334
Household member reported cough, fever, shortness of breath, abnormal taste/smell
n= 160
n= 1174
Anti-Spike or anti-N immunoassays > 0.35 (possible antibody responses)
n= 87
Low SARS-CoV-2 risk group n= 1087
674 females, 413 male
Key workers with previous RT-PCR test positive
Prior positive group
n= 268
188 females, 80 males
High risk
Low risk
Anti-Spike IgG confirmed positive (EuroImmun index >= 1.1)
Anti-Spike IgG not confirmed positive(EuroImmun index < 1.1)
B
SNM T-SPOT result (count/250,000 cells)
300
100
30
10
5
4
3
2
1
0
Low risk
Prior Positive SARS-CoV-2 RT-PCR
Participant category
C
anti-S immunoassay signal (EuroImmun)
10.0
1.0
0.1
Low risk
Prior Positive SARS-CoV-2 RT-PCR
Participant category
D
anti-S immunoassay signal (EuroImmun)
10.0
1.0
0.1
0
1
2
3
4
5
10
30
100
300
SNM T-SPOT results (count/250,000 cells)

Fig. S7

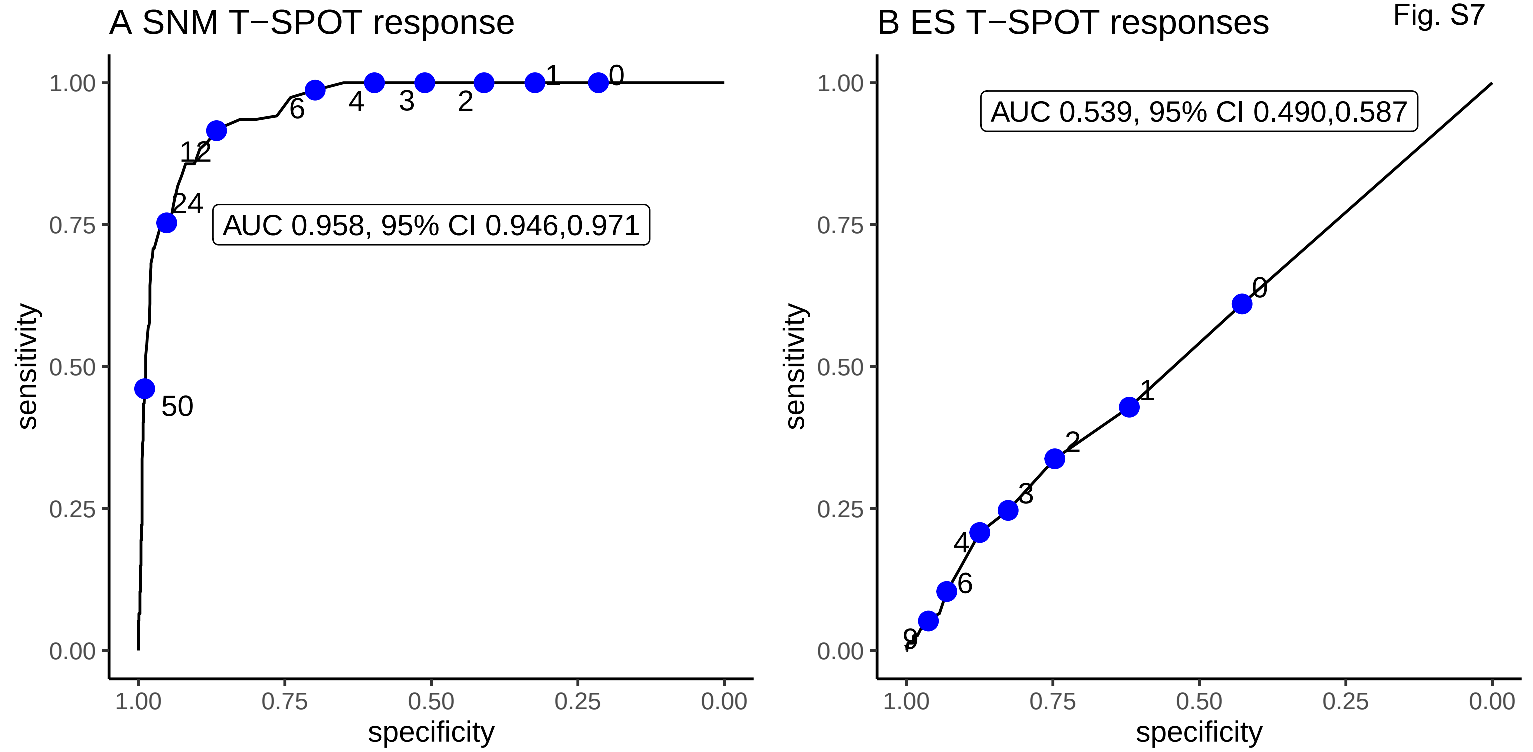
A SNM T−SPOT response
AUC 0.958, 95% CI 0.946,0.971
sensitivity
specificity
1.00
0.75
0.50
0.25
0.00
50
24
12
6
4
3
2
1
0
B ES T−SPOT responses
AUC 0.539, 95% CI 0.490,0.587
sensitivity
specificity
9
6
4
3
2
1
0
